# Integrated real-world and translational studies nominate CREB binding protein/EP300 inhibition to overcome androgen receptor pathway modulator resistance in men with prostate cancer

**DOI:** 10.64898/2026.11.24.26368506

**Authors:** Jason A. Somarelli, Bonnie V. Dougherty, Aviva G. Beckmann, Ananya Dutta, Beatrice C. Thomas, Alice M. Walsh, Duo Xu, Veronica Calvo Vidal, Pelumi D. Olawuni, Maureen Caligiuri, Erik J. Soderblom, Sylvie M. Guichard, Phillip Comella, Seb Monzon, Andre N. Forbes, Jens Renstrup, Eric E. Schadt, Andrew J. Armstrong

## Abstract

Androgen pathway modulators (APMs) have significantly improved outcomes for men with metastatic prostate cancer, but the evolution of therapy resistance is nearly universal, and there remains a major clinical need to identify actionable vulnerabilities for therapy-resistant disease. To address this need, we employed regulatory network models that leverage multi-omic data from over 10,000 prostate cancer patients with longitudinal clinical data. Analysis of the architecture of the network models identified EP300 as the top actionable target associated with androgen receptor-mediated therapy resistance. These observations were validated using a novel inhibitor of CBP/EP300, pocenbrodib, which showed preclinical activity *in vitro* and *in vivo*. Acetylomics analysis and functional studies pinpointed inhibition of oxidative phosphorylation and fatty acid metabolism as key mechanisms of action for pocenbrodib. Clinical benefit for this agent was demonstrated in a patient with metastatic APM-resistant prostate cancer (NCT04575766), highlighting CBP/EP300 as a key target for further translation.

## Introduction

Despite the successful development of DNA-level precision oncology biomarkers in men with prostate cancer, there have been limited successes so far in integrating transcriptomics or other multi-omics biomarkers into clinical decision-making (1-3). One of the limitations in broadly translating precision oncology strategies from research discovery to the clinic is the identification and clinical validation of biomarkers that are capable of pinpointing actionable molecular mechanisms most responsible for variations in clinical outcomes. While numerous prognostic omics-based signatures have been developed to stratify the prognosis of patients by clinical outcomes (4), today there are still no predictive complex, multi-omic biomarkers appearing on the label of any marketed cancer therapeutic, given these signatures have been unable to decipher core master regulators operating within gene regulatory networks that promote a given phenotype associated with outcomes, such as overall or progression-free survival.

The lack of ability to identify key master regulators, however, is becoming increasingly addressable as the scales of real-world, multimodal data generated on cancer patients in current-day standard-of-care settings continue to grow rapidly. This includes comprehensive, multi-omics data that is matched to longitudinal clinical data, enabling the reconstruction of increasingly-accurate patient journeys. Such data can provide the density and longitudinal scales needed to identify and directly link mechanisms that are driving resistance to standard-of-care therapeutic interventions, and to match these mechanisms to targets that may be suitable for therapeutic development. The critical need for advancements in this domain is particularly evident in prostate cancer, which ranks as the second leading cause of cancer-related death in the United States (5). In this context, addressing the challenge of metastatic and therapy-resistant disease, the major causes of death for cancer patients, becomes not only urgent, but also a potential area for groundbreaking therapeutic developments.

The mainstay of treatment for patients with metastatic prostate cancer is inhibition of the androgen receptor (AR) signaling pathway through androgen deprivation therapy (ADT) and targeted AR pathway modulators (APMs) (6) or anti-androgens, which are collectively referred to as hormone therapy. These therapies have led to significant improvements in survival and have become the standard-of-care for patients with metastatic APM sensitive/naïve or -resistant prostate cancers (7-13). Despite the benefit of these treatments, however, resistance to AR-targeted therapy is nearly universal; patients develop progressive, hormone-therapy resistant disease over time, which has a poor prognosis and few treatment options.

To better understand disease progression of current-day patients with prostate cancer as they transition to hormone therapies and develop resistance to such therapies we employed a real-world data-driven approach to elucidate mechanisms of resistance and identify therapeutic targets capable of addressing these mechanisms through alternative means, independently of directly targeting AR. Utilizing a cohort of 12,296 prostate cancer patients with longitudinal clinical record data matched to tumor DNA and whole transcriptome RNA sequencing data, we constructed comprehensive regulatory network models in late-stage prostate cancer. These analyses revealed subnetworks linked to cell cycle, E2F, MYC, AR signaling, and oxidative phosphorylation that were most significantly associated with disease progression and resistance to hormone therapies in our cohort. By analyzing the architecture of our regulatory network models of prostate cancer, we identified EP300 as the top actionable target underlying the driver networks mechanisms associated with resistance and progression of prostate cancer.

To empirically validate the real-world data insights our models revealed, we employed a novel small-molecule inhibitor of CBP/EP300, pocenbrodib, in active clinical development (14), targeting the CBP/P300 bromodomain. CBP/EP300 inhibition significantly reduced proliferation of all prostate cancer cell lines and demonstrated *in vivo* efficacy in patient-derived xenografts. Further, administration of pocenbrodib demonstrated the potential for clinical benefits and durable responses in the COURAGE clinical trial (NCT04575766) for metastatic, APMR prostate cancer. Molecular profiling of cell lines pre- and post-CBP/p300 inhibitor treatment demonstrated inhibition of the core molecular mechanisms identified in the network models from patients. Proteomics and acetylomics from enzalutamide-sensitive and -resistant LNCaP95 cells revealed global reprogramming of the acetylome, with significant differential acetylation and expression of proteins involved in oxidative phosphorylation and fatty acid metabolism, as predicted by the regulatory networks. Consistent with this, CBP/EP300 inhibition led to inhibition of oxidative phosphorylation, a reduction in acetyl CoA pools and loss of lipid droplets, suggesting inhibition of oxidative phosphorylation and fatty acid oxidation as key mechanisms of CBP/EP300 inhibition. These observations highlight CBP/EP300 as a key target for further clinical translation in patients with hormone therapy-resistant prostate cancer.

## RESULTS

### Characterizing the advanced prostate cancer treatment journey in a large real-world dataset: from diagnosis to castration resistance

We assembled a cohort of 12,296 prostate cancer patients that included DNA sequencing and/or RNA sequencing from isolated tumor tissue paired with longitudinal clinical record data (**Figure 1A**). Samples that met inclusion criteria with available sequencing and clinical data were stratified by disease setting, resulting in 1,971 and 718 samples from patients with metastatic, APM sensitive or naïve and APM resistant, respectively, available for downstream analysis (**Figure 1A**). Within this cohort, we observed that ADT monotherapy and ADT + taxane regimens were increasingly replaced by androgen receptor pathway inhibitor-containing doublet and triplet as first-line therapy from 2018 onward (**Figure 1B**). This shift reflects the sequential FDA approvals of abiraterone, apalutamide, enzalutamide, and darolutamide in the mHSPC setting, supported by the landmark ARCHES, ENZAMET, ARASENS, and ARANOTE trials (8, 12, 15-18). In our data, first-line treatment intensification beyond ADT monotherapy was associated with improved OS from the time of first therapy (global log-rank p=0.0013; **Figure 1C, D**). Compared to ADT alone (median OS 30.3 months), both ADT + APM doublet therapy (median OS 36.7 months; HR 0.76, 95% CI 0.63–0.92) and ADT + APM + taxane triplet therapy (median OS 41.5 months; HR 0.60, 95% CI 0.45–0.81) demonstrated significantly improved OS, with greater benefit observed with triplet therapy. Notably, these survival differences were observed in the absence of adjustment for known confounders of treatment selection in real-world settings, such as disease volume, performance status, and comorbidity burden, suggesting that the OS benefit associated with APM-based intensification is robust and likely conservative relative to what would be observed in a covariate-adjusted analysis. Reflecting this shift in treatment paradigm towards earlier treatment intensification, the proportion of patients receiving their first APM in the mHSPC setting has increased substantially over time; in 2024, 80% of patients had previously been exposed to a prior APM at the time of APM resistant diagnosis (**Supplemental Figure 1**).

**Figure 1.**
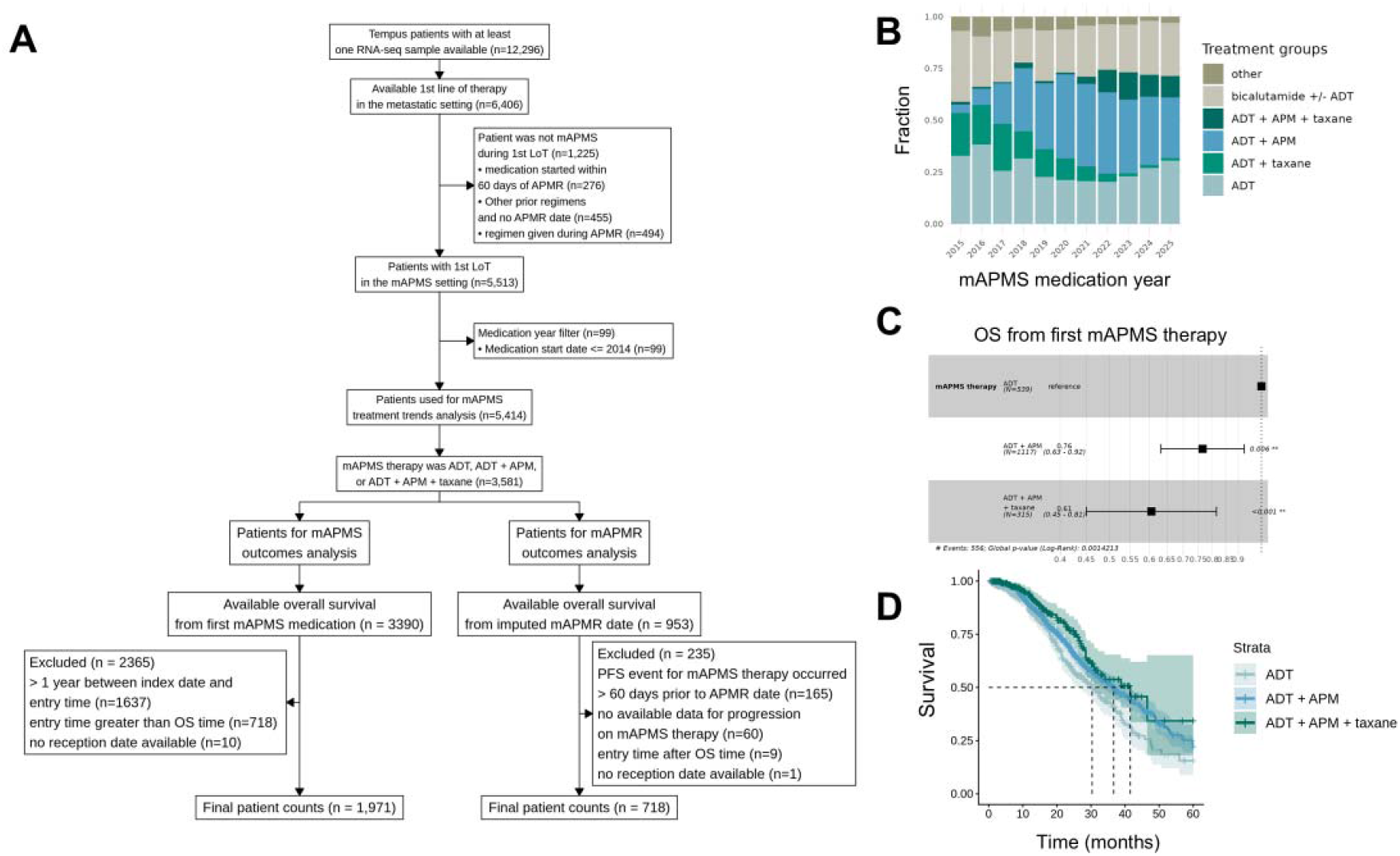
Androgen pathway modulator-based treatment intensification in the metastatic androgen pathway modulator sensitive (APMS) setting is associated with better overall survival. (A) CONSORT diagram depicting patient selection from the Tempus real-world dataset which required at least tissue sample with available DNA or RNA data. (B) Treatment landscape trends from 2015 to 2025 demonstrate the progressive displacement of ADT monotherapy by APM-containing doublet and triplet regimens in the first-line setting in mAPMS disease. (C) Real-world overall survival from the start of first APMS therapy demonstrates a significant and stepwise improvement with treatment intensification, with triplet therapy showing the greatest OS benefit relative to ADT monotherapy. (D) Real-world overall survival from the start of mAPMS therapy reveals patients treated with triplet therapy have a higher median OS of 10 months compared to patients treated with ADT monotherapy.

From our cohort, we next analyzed a subset of 12,608 samples from the Tempus real-world database with available tumor DNA and/or RNA profiling of tumor tissue, comprising 8,977 samples collected in the metastatic, androgen pathway modulator sensitive (mAPMS) setting and 1,142 samples collected in the metastatic, androgen pathway modulator resistant (mAPMR) setting (**Figure 2A**). Comparison of the genomic landscape across these disease states revealed substantial differences in the prevalence and spectrum of somatic alterations between disease states (**Figure 2B**). While alterations in TP53 and PTEN were present across both settings, their frequency appears to increase in mAPMR patient tumors compared to hormone naïve or responsive tumors. In contrast, AR, including both mutations and copy number gains as well as RB1 alterations appear frequently in patients with mAPMR but not in mAPMN/S disease. To better understand the emergence of APM resistance mechanisms and account for potential treatment confounders, such as chemotherapy, we restricted subsequent analyses to samples collected after progression on an APM, but prior to initiation of non-AR-based therapy (**Figure 2C**, n=544). To identify genomic alterations enriched in the post-APM failure setting, we performed Fisher’s exact tests comparing alteration frequency between pre-APM (n=1,246) and post-APM failure (n=544) samples across the top 10 most frequently altered genes (**Figure 2C**; FDR < 0.05). AR and RB1 alterations were the most significantly enriched in the post-APM failure setting compared to the pre-APM setting (**Figure 2D**) (19, 20). Given the complexity of the post-APM genomic landscape, we next examined pairwise co-occurrence of the most frequently occurring alterations, AR, RB1, PTEN, and TP53 within the post-APM failure samples (FDR < 0.05; **Figure 2E**). TP53 and RB1 alterations demonstrated significant co-occurrence (log2 OR = 2.58; FDR = 0.003; **Figure 2E**), consistent with their established losses in advanced prostate cancer that promote neuroendocrine-like lineage reprogramming (21, 22). In contrast, RB1 and AR alterations were significantly mutually exclusive (log2 OR = 0.43; FDR = 0.03) (**Figure 2E**), suggesting that AR and RB1 alterations represent distinct and non-overlapping mechanisms of APM resistance.

**Figure 2.**
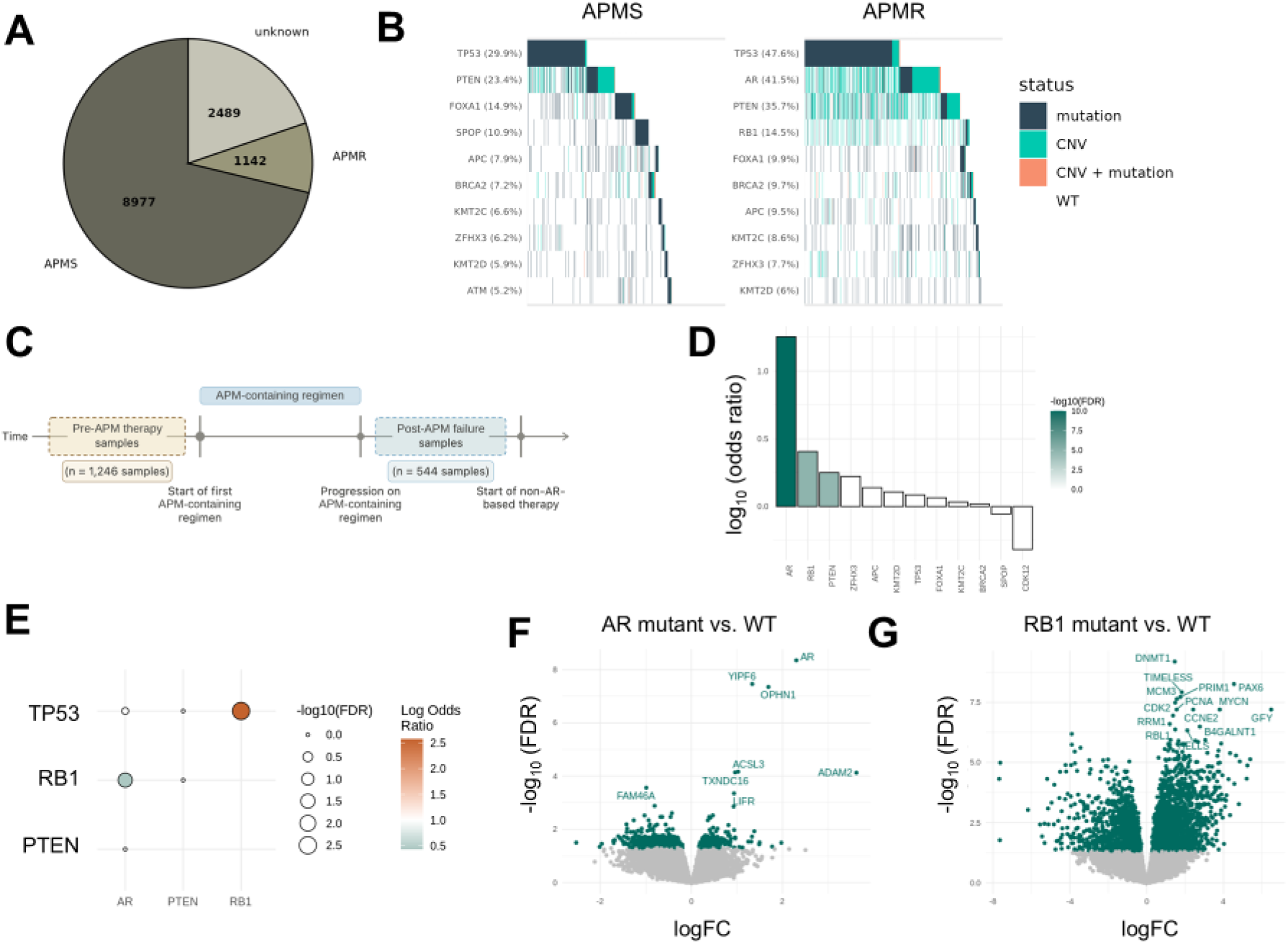
Genomic and transcriptomic characterization of APM resistance mechanisms in real-world prostate cancer samples. (**A**) Distribution of available tissue biopsies across disease settings. (**B**) Oncoprint depicting the most frequent DNA alterations in the APMS (left) and APMR (right) samples, highlighting the emergence of AR amplifications, AR mutations, and RB1 loss in APMR samples. (**C**) Schematic depicting the sample selection strategy for pinpointing mechanisms of resistance to APM therapies, with pre-APM samples (n = 1,246) collected prior to the start of the first APM-containing regimen and post-APM failure samples (n=544) collected following progression on an APM-containing therapy but prior to the start of a non-AR based therapy. (**D**) Enrichment of genomic alterations in the post-APM failure samples relative to the pre-APM samples demonstrates significant enrichment for AR, RB1, and PTEN alterations in the post-APM setting. (**E**) Co-occurrence analysis of key genomic alterations demonstrates the co-occurrence of TP53 and RB1 alterations and the mutual exclusivity of AR and RB1 alterations. (**F**) Volcano plot of differential expressed genes in the AR altered vs AR WT samples highlights upregulation of AR and ASCL3. (**G**) Volcano plot of differentially expressed genes in the RB1 altered vs RB1 WT samples highlights broad up regulation of cell cycle and neuroendocrine associated genes.

To identify the downstream molecular mechanisms unique to each resistance mutation while accounting for co-occurrence of RB1 and TP53 and mutual exclusivity of RB1 and AR alterations, we performed differential expression analyses on tumors harboring AR alterations or AR WT without co-occurring RB1, TP53, or PTEN alterations (**Figure 2F**) and tumors harboring RB1 alterations or RB1 WT without co-occurring AR, TP53, or PTEN alterations (**Figure 2G**). In AR-altered tumors, AR was the most significantly upregulated gene alongside two genes involved in fatty acid metabolism, LIFR and ASCL3 (**Figure 2F**). Gene set enrichment analysis (GSEA) of the AR signature demonstrated significant upregulation of the Hallmark androgen response, MYC, oxidative phosphorylation and E2F gene sets (**Supplemental Figure 2A**). In contrast, the RB1 altered signature was characterized by coordinated upregulation of canonical E2F and cell cycle genes (e.g., CDK2, CCNE2, PCNA, MCM3, and MYCN; **Figure 2F)** and associated pathways (**Supplemental Figure 2B**). These observations are consistent with activation of a lineage plasticity program consistent with neuroendocrine differentiation upon RB1 loss in prostate cancer (22-24).

### Regulatory networks identify master regulators associated with resistance to androgen pathway modulators in prostate cancer patients

To identify novel actionable targets for patients with APM-resistant prostate cancer we next analyzed samples collected after the start of the first APM regimen to construct comprehensive regulatory networks that synthesize diverse data types including gene expression, single nucleotide variants (SNVs), copy number variations (CNVs), and clinical features, using a multi-network construction framework that integrates co-expression, causal mediation, and probabilistic causal network construction methodologies (**Figure 3A**). Using weighted gene co-expression network analysis (WGCNA) we identified 40 subnetworks with > 30 highly co-regulated genes from the post-APM samples. Clustering from the eigengene pinpointed 13 superclusters among these co-expression modules (**Figure 3B**). Hallmark pathways most significantly represented across the superclusters included immune and inflammatory signaling (M1), proliferation and cell cycle (M3), MYC targets (M5, M6, and M9), hormone response (M7), tissue identity / differentiation (M2), and metabolism (M4, M5), representing major programs that are associated with prostate cancer (**Figure 3B**).

**Figure 3.**
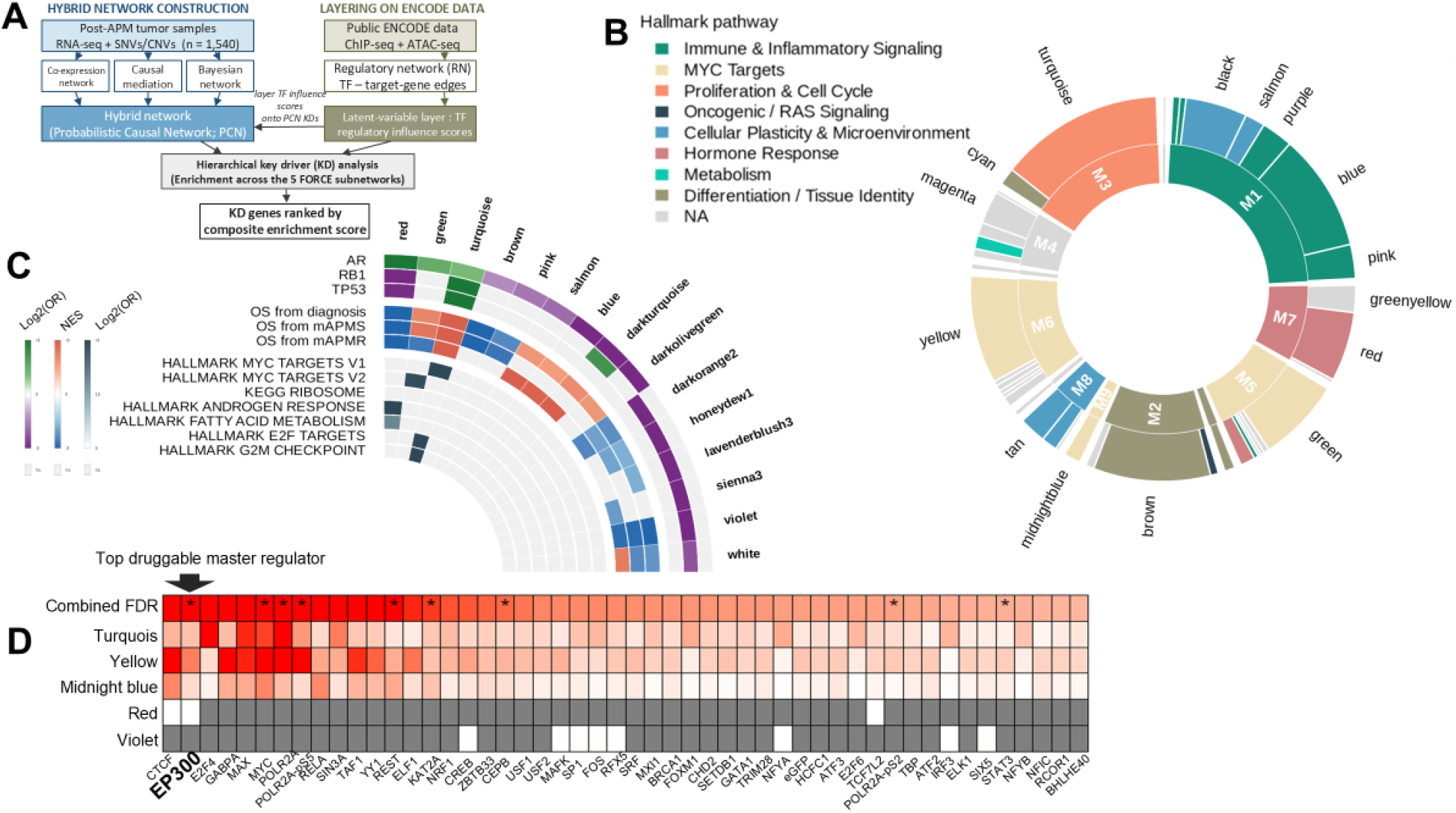
Co-expression network analysis identifies APM resistance-associated transcriptional subnetworks converging on EP300 as a therapeutic target. (**A**) Schematic depicting the sample selection strategy for co-expression network analysis, with pre-treatment samples (n=2,634) collected prior to the start of any therapy and post-APM samples (n=1,540) collected following initiation of the first APM-containing regimen. (**B**) Donut plot depicting the functional annotation of WGCNA superclusters and subnetworks by Hallmark pathway category, with major superclusters (M1-M8) capturing distinct biological programs including proliferation and cell cycle, immune and inflammatory signaling, metabolism, oncogenic signaling, and hormone response. (**C**) Circos plot depicting the association of WGCNA subnetworks with AR and RB1 alteration signatures, overall survival from diagnosis, mAPMS, and mAPMR, and Hallmark pathway enrichment. (**D**) Validation of AR signature-associated subnetworks using publicly available RNA-seq data from LNCaP and VCaP cell lines treated with varying concentrations of the synthetic androgen, R1881. (**E**) ENCODE ChIP-seq enrichment analysis identifying transcription factors whose target genes are broadly enriched across the five AR signature-associated subnetworks, with EP300 emerging as the top-ranked candidate showing FDR-significant enrichment across the identified subnetworks, supporting its prioritization as a therapeutic target for disrupting resistance-associated transcriptional programs.

To further characterize the downstream transcriptional consequences of genomic alterations associated with APM resistance, we overlaid the signatures of AR, RB1, and TP53 alterations onto the co-expression network using Fisher’s exact test to identify subnetworks significantly enriched for each alteration-associated transcriptional program. The up-regulated genes in the AR signature were significantly enriched in 5 subnetworks, representing canonical AR signaling, MYC signaling, and cell cycle progression (**Figure 3C**), reflecting the breadth of transcriptional reprogramming driven by AR pathway activation in late-stage prostate cancer. In addition, the red subnetwork was significantly enriched for the Hallmark androgen response as well as the Hallmark fatty acid metabolism gene set, suggesting that AR and fatty acid metabolism are not only co-activated in AR-altered tumors, but are transcriptionally coordinated (**Figure 3C**).

The RB1 signature was significantly enriched in the two subnetworks (turquoise and cyan) belonging to the M1 supercluster (**Figure 3C**), which was enriched for cell cycle related genes, consistent with the established role of RB1 loss in promoting lineage plasticity and neuroendocrine plasticity. Notably, the AR and RB1 signatures showed distinct and largely non-overlapping module enrichment patterns, consistent with their representing complementary transcriptional axes of APM resistance. However, despite their distinct primary enrichment patterns, the AR and RB1 signatures showed overlapping enrichment in the proliferation and cell cycle subnetwork (**Figure 3C**; turquoise), demonstrating that despite distinct primary enrichment patterns, AR activity, RB1 loss, and TP53 alterations converge on transcriptional activation of E2F-driven proliferative programs.

To understand whether the identified co-expression subnetworks are associated with patient outcomes, we used GSEA to project survival-ranked gene signatures derived from overall survival analyses across three disease contexts: diagnosis, APMS, and APMR (**Figure 3C**). Subnetworks enriched for survival-associated gene programs were largely consistent across all three disease contexts (**Figure 3C**). Of note, the subnetwork enriched for the Hallmark androgen response is also enriched in genes related to oxidative phosphorylation and fatty acid metabolism, suggesting potential altered metabolic states during APM progression.

To further resolve whether the identified subnetworks may be associated with AR activity in the clinical data, we performed Fisher’s exact tests comparing subnetworks with public RNA-seq data from two cell lines (LNCaP and VCaP) treated with various concentrations of synthetic androgen, R1881 (25). The subnetworks that capture AR signaling genes (turquoise, red, and midnight blue, green, and brown) were the top subnetworks enriched for the R1881 stimulation differentially expressed genes, further supporting that these subnetworks capture AR-driven transcriptional programs (**Supplemental Figure 2C**). Interestingly, the subnetwork enriched in MYC signaling (yellow), was ranked lower than a separate subnetwork that also captures MYC signaling (green; **Supplemental Figure 2C**), suggesting that the MYC-associated transcriptional program captured in patient tumors is not fully recapitulated by androgen stimulation alone. More broadly, the direction and magnitude of the subnetwork associations with R1881 were not consistent across concentrations or cell lines, suggesting that AR alterations in the context of advanced disease engage and re-wire AR-associated transcriptional programs beyond what is observed under androgen simulation *in vitro* (**Supplemental Figure 2C**). Overall, this approach identified five subnetworks that capture genes up regulated in AR altered samples that are also associated with clinical outcomes across the spectrum of disease. We refer to these subnetworks as the functional outcomes in real-world cancer environments (FORCE) subnetworks and hypothesize that targeting these subnetworks therapeutically could not only address mechanisms of resistance to APMs, but also delay or prevent the development of resistance.

To identify regulatory targets that modulate these subnetworks, we developed a hierarchical network strategy that involves the construction of two complementary network types: 1) probabilistic causal network (PCN) models to explicitly represent inferred causal relationships among nodes (26-32); and 2) regulatory networks (RNs) constructed by integrating ChIP-seq and ATAC-seq data generated in independent, context-relevant datasets. In this analysis, key driver genes are defined as network nodes that are predicted to modulate significantly enriched nodes within any of the 5 FORCE subnetworks. The full list of key driver genes identified in this analysis is given in **Supplementary Table 1**.

To identify transcription factors whose target genes are broadly enriched across the 5 FORCE subnetworks, we performed a gene set enrichment analyses using ENCODE ChIP-seq data and ranked TFs and combined their respective p-values across these subnetworks (**Figure 3D**). The violet subnetwork (**Figure 3D**, number 4) yielded only one significant enrichment in one cell line, most likely due to the subnetwork representing a small number of genes (n = 42). The identified TFs were then cross-checked with available clinically-actionable agents to prioritize targets for further study (**Figure 3D**, asterisks). Among all TFs tested, EP300 was the top-ranked actionable candidate showing FDR-significant enrichment across the turquoise, yellow, midnight blue, and red subnetworks, supporting its candidacy as a broadly-relevant therapeutic target across all five resistance-associated transcriptional programs (**Figure 3D**).

### CBP/EP300 inhibition is efficacious in preclinical models of APM-resistant prostate cancer

To test the relevance of EP300 in prostate cancer, we evaluated the preclinical efficacy of a novel CREB binding protein/EP300 inhibitor, FT-6876 (a tool compound for pocenbrodib) in a panel of five prostate cancer cell lines that represent a spectrum of AR signaling dependency, including the AR+ LNCaP, VCaP, and 22Rv1, and AR- PC-3 and DU145 cells. As predicted by the enrichment of EP300 in AR-driven FORCE subnetworks, we observed greater cell growth inhibition in the AR+ cell lines (**Figure 4A**). We also tested the capacity of CBP/EP300 inhibition to suppress the growth of enzalutamide-resistant disease using three isogenic pairs of enzalutamide-sensitive and –resistant sub-lines, LNCaP, LNCaP-CS2, and LNCaP95, all of which showed dose-dependent reductions in growth rate after CBP/EP300 inhibition (**Figure 4B**). CBP/EP300 inhibition also significantly reduced colony forming capacity (**Figure 4C**) and sphere formation capacity (**Figure 4D**) in LNCaP95 enzaS and enzaR lines. Similar results were observed in LNCaP-CS2 enzaS and enzaR lines and in CWR1 cells engineered with the AR truncated ligand receptor variant, ARv567es, which renders cells resistant to enzalutamide (**Supplemental Figure 3**). We also observed efficacy of CBP/EP300 inhibition in the AR+ enzalutamide-resistant patient-derived xenograft (PDX), CTG-2440 (**Figure 4E**) and two additional PDXs, CTG-2430 (wtAR, Enza-sensitive) and CTG-2428 (wtAR, Enza-sensitive; **Supplemental Figure 3**).

**Figure 4.**
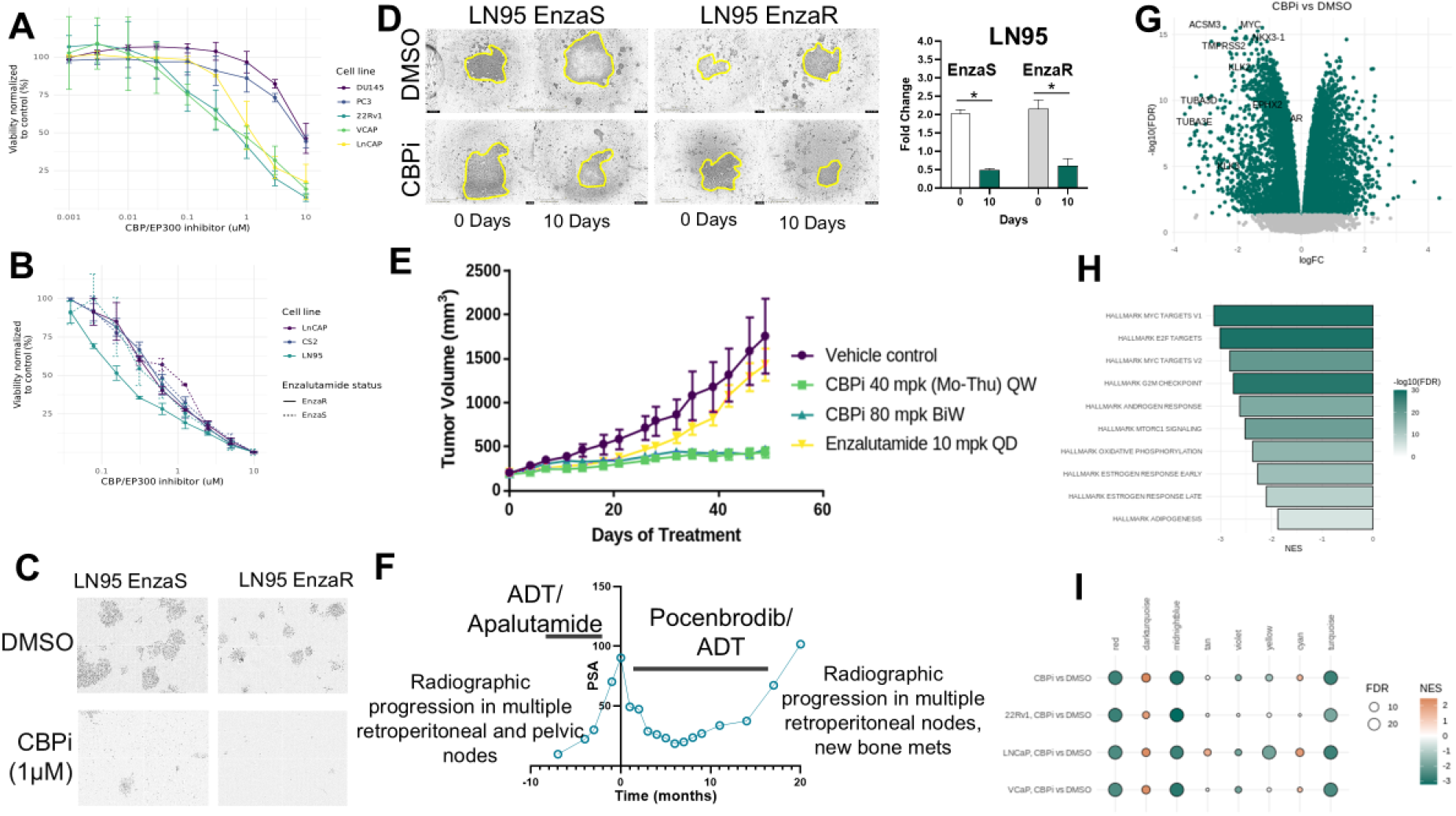
Efficacy of CBP/EP300 inhibition in APM-resistant prostate cancer. **A.** Dose-response curves for 5 prostate cancer cell lines. **B.** Dose response curves for isogenic pairs of enzalutamide-sensitive and –resistant lines. **C.** Colony forming assays in DMSO- and CBPi-treated LNCaP95 cells. **D.** Spheroid forming assays for LNCaP95 enzalutamide-sensitive and –resistant spheroids treated with vehicle (DMSO) or CPBi. **E.** *In vivo* evaluation of CBPi in an enzalutamide-resistant PDX. **F.** PSA dynamics for patient treated with pocenbrodib. **G.** Volcano plot of differentially-expressed mRNAs in LNCaP, 22Rv1, and VCAP quantified by RNA-Seq. **H.** GSEA pathway analysis from RNA-Seq on LNCaP, 22Rv1, and VCAP cells treated with vehicle (DMSO) or pocenbrodib (CBPi). **I.** Enrichment in the differential expression signatures for each of these lines across the FORCE subnetworks

The promising preclinical activity of CBP/EP300 inhibition led to an open-label phase I dose escalation trial (NCT04575766; lead compound pocenbrodib was used in the human study) with the primary outcome to evaluate the safety and tolerability of this novel inhibitor in men with mAPMR disease. Secondary outcomes included evaluating evidence of clinical activity as defined by PSA declines, radiographic responses, and radiographic progression free survival, defined by PCWG3 and RECIST 1.1 criteria. In this phase 1 trial, clinical benefit from pocenbrodib was observed for a man in his 60s with relapsed mAPMR prostate cancer. This patient developed disease progression despite prior surgery, radiation, ADT, abiraterone, and apalutamide. The PSA on apalutamide declined but rose to 70 by early 2022, at which time metastatic adenopathy developed on CT imaging indicating the patient had developed metastatic CRPC/APMR. The patient elected to defer taxane chemotherapy and consented to the Courage phase 1 study of FT-7051 (NCT04575766). Cycle 1 was initiated while PSA was 90 (**Figure 4F**). The patient was treated with pocenbrodib on an intermittent dosing schedule within 28-day cycles, starting at a dose within the range evaluated in the trial. PSA was measured prior to each cycle and the patient underwent CT and bone scan every other cycle. His PSA decreased from 90 prior to initiation of the trial to 19.9 (78% PSA decline) after 7 cycles of therapy (**Figure 4F**). Imaging after cycles 2, 4, and 6 did not show any evidence of metastatic disease and the retrocrural 1.3 cm node that was detected prior to initiation remained stable in size. After 20 months of therapy, he developed consecutive PSA rises and radiographic progression in lymph node and new bone metastases and came off trial. Over the course of treatment the patient did not experience any grade 4 adverse effects (AEs). Dose reduction was required after several cycles due to treatment-related adverse events, including grade 3 fatigue and grade 2 hyperglycemia, and the patient was subsequently maintained at a lower, well-tolerated dose. Reported AEs at this dose included grade 1 nausea and lipase elevation, and grade 2 dysgeusia and dyspnea on exertion. Pocenbrodib is being further evaluated clinically in the PATHWAY clinical trial (NCT06785636). Together, these data suggest that CBP/P300 inhibition has the potential to overcome a range of AR-dependent mechanisms of APM resistance in prostate cancer.

To infer the underlying mechanisms of action for the CBP/EP300 inhibitor, we sequenced RNA isolated from LNCaP, 22Rv1, and VCAP prostate cancer cell lines after 24 hours (1μM) and observed significant downregulation in multiple FORCE subnetworks related to AR-mediated mechanisms of APM resistance, including Myc, androgen response, and oxidative phosphorylation (**Figure 4G, H**). Similarly, we observed enrichment in the differential expression signatures for each of these lines across the FORCE subnetworks of interest, as predicted by our model (**Figure 4I**). In all cases the modules were statistically significantly enriched (p < 1e-15) in at least three of the prostate cancer models, and all of the subnetworks were enriched for the composite signature, generated by identifying differentially expressed genes between the combined treated and untreated cell lines (**Figure 4I**).

### CBP/EP300 inhibition rewires the acetylome of key metabolism genes

Given the regulatory impact of CBP/P300 inhibition on the key driver mechanisms predicting outcomes in the post-APM setting, and the established function of CBP/P300 as an acetyltransferase, we sought to characterize the consequences of CBP/P300 inhibition on the downstream proteome and acetylome of prostate cancer cells. To do this, we performed global proteomics and acetylomics on LNCaP95 enzalutamide-sensitive and -resistant cells treated with either vehicle or CBP/EP300 inhibition (1 μM) for 24 hours (**Figure 5A**). Western blotting for H3K27ac confirmed inhibition of the histone acetyltransferase activity of CBP/P300 (**Figure 5B**). At FDR<0.05, analysis of differential protein abundance resulted in the identification of 241 upregulated and 252 downregulated proteins in enzalutamide-sensitive LNCaP95 cells, and 178 upregulated and 240 downregulated proteins in enzalutamide-resistant LNCaP95 cells (**Figure 5C**). Of these proteins, eight (NCDN, RASA1, GRB10, RMDN3, PC, MED7, IDH3G, PIN4) were commonly upregulated and six (CBWD3, ATAD1, YRDC, DGKH, DHTKD1, FLVCR1) were commonly downregulated between CBP/P300i-treated enzalutamide-sensitive and -resistant cells. From the acetylomics data we identified 253 differentially acetylated proteins in enzalutamide-sensitive LNCaP95 cells and 254 in enzalutamide-resistant cells (**Figure 5D**). Using principal component analysis of proteomic profiles we observed CBP/p300 inhibition in enzalutamide-resistant cells shifts the proteomic state towards the untreated enzalutamide-sensitive state along PC2 (**Figure 5E**), consistent with the potential partial reversal of the resistance-associated proteome. At the pathway level, differentially acetylated proteins were significantly enriched in ribosomal pathways, Myc, and oxidative phosphorylation (**Figure 5F**), with MYC also identified as a key regulator of the 5 FORCE subnetworks (**Figure 3F**). Overlapping acetylated proteins included multiple proteins involved in epigenetic remodeling (histones), fatty acid metabolism, and oxidative phosphorylation (**Figure 5G, H**). Comparison of TOM values for AR versus key genes in the androgen response or fatty acid metabolic pathways highlights connectivity between AR and fatty acid metabolism sub-networks in the post-treatment networks not present in the pre-treatment networks (**Figure 5I**).

**Figure 5.**
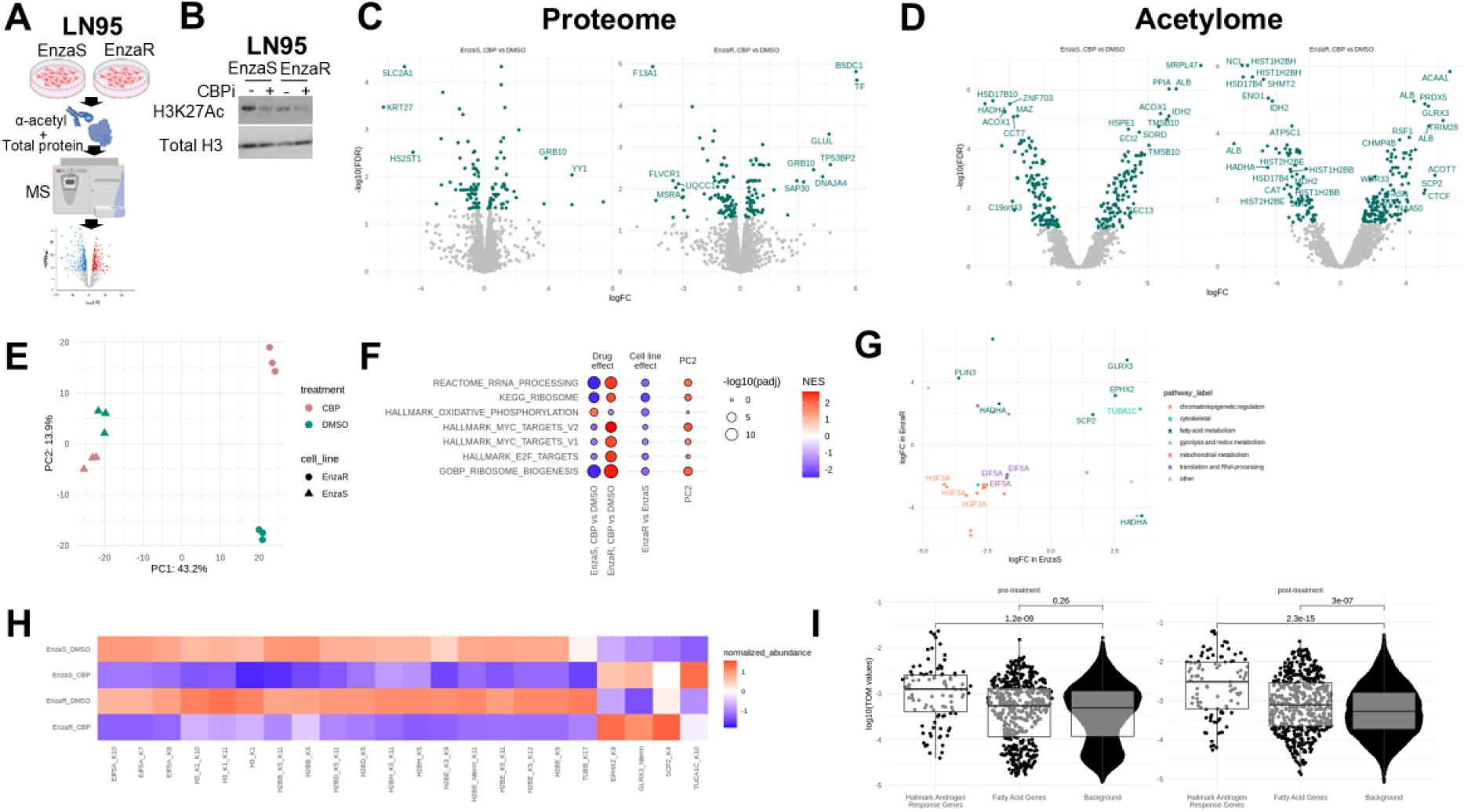
Acetylomics identifies differential acetylation of key oxidative phosphorylation and fatty acid metabolism genes. (**A**) Schematic workflow for proteomics and acetylomics analysis. (B) Western blot for H3K27Ac. Total histone H3 levels serve as a loading control. (**C**) Volcano plot for differentially abundant proteins between the enzalutamide-sensitive and -resistant cell lines. (D). Volcano plot for differentially acetylated proteins in enzalutamide-sensitive and –resistant cells. (**E**) Principal component analysis of proteomic profiles reveals that CBP/p300 inhibition in the enzalutamide-resistant line shifts the proteomic state towards the enzalutamide-sensitive (EnzaS) along PC2, consistent with the potential partial reversal of the resistance-associated proteome. (**F**) Pathway enrichment of differential protein abundance. (**G**) Scatter plot of acetylation sites that are significantly differentially acetylated in the enzalutamide-sensitive and –resistant cells following treatment with CBPi, highlighting key shared changes in fatty acid metabolism. (**H**) Heatmap of normalized abundance changes for the acetylation sites that are shared in direction and significance in enzalutamide-sensitive and –resistant cells. (**I**) Comparison of TOM values for AR versus key genes in the androgen response or fatty acid metabolic pathways highlights correlation structure between AR and fatty acid metabolism in the post-treatment networks that is not present in the pre-treatment networks.

Our systems-level analysis of patient samples and preclinical models pinpointed alterations in metabolic axes (oxidative phosphorylation, fatty acid metabolism) as key features of APM resistance that are potentially targetable via CBP/EP300 inhibition. To experimentally validate these metabolic alterations, we performed Seahorse analysis to quantify changes in oxidative phosphorylation during pharmacological inhibition of CBP/EP300. Treatment with CBP/EP300 inhibitor led to significant reduction in oxidative phosphorylation (**Figure 6A, B**). The reduction in oxidative phosphorylation was accompanied by significant loss of acetyl-coA levels (**Figure 6C, D**) and a reduction in lipid droplet accumulation (**Figure 6E, F**). These data suggest that the epigenetic remodeling of the prostate cancer proteome and acetylome by CBP/EP300 inhibition leads to a profound metabolic suppression, which contributes to its anticancer activity.

**Figure 6:**
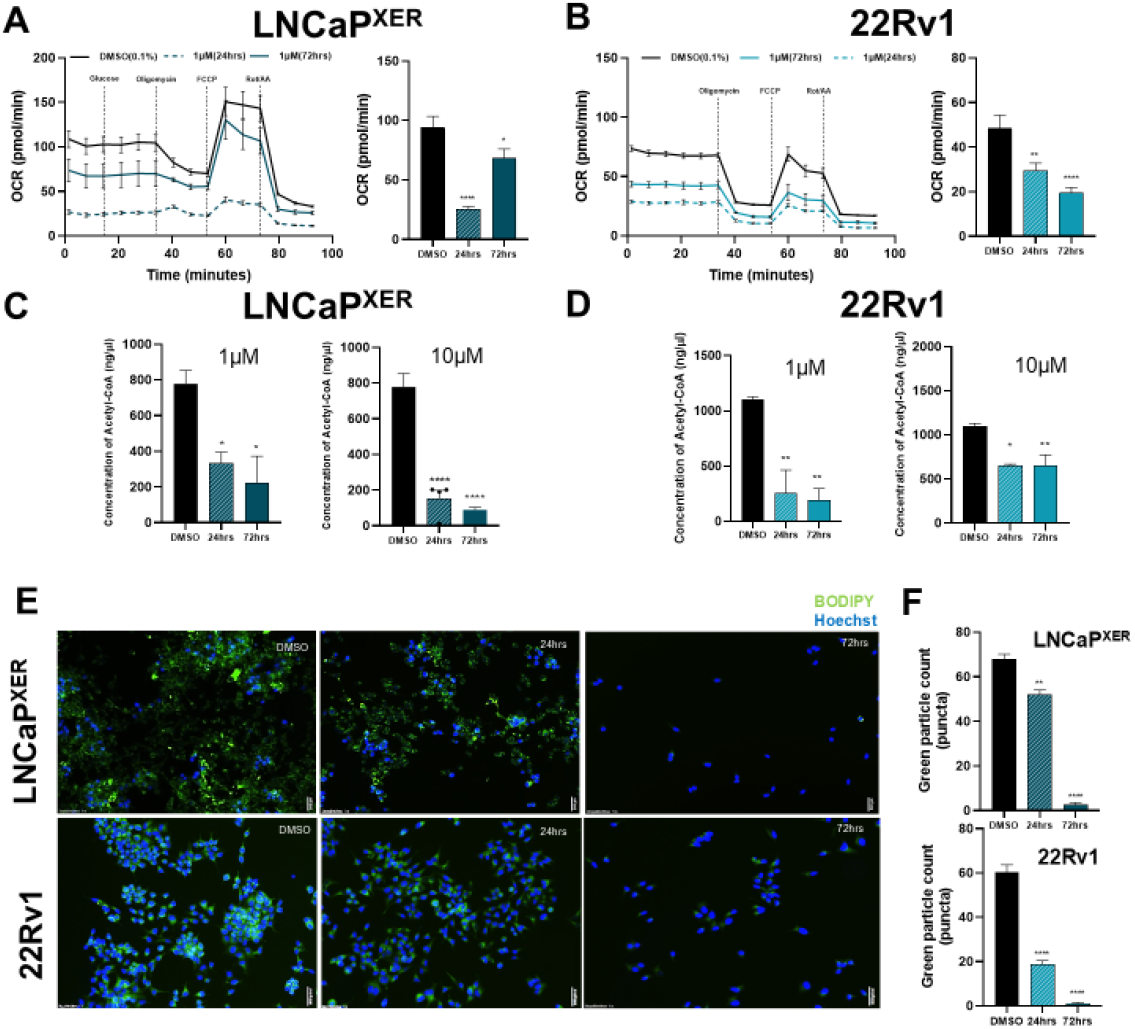
Pocenbrodib-mediated CBP inhibition impairs oxidative phosphorylation. **A.** Oxygen consumption rate as assessed by Seahorse XF analysis at 1 and 10 µM treatment with pocenbrodib in LNCaP^XER^ and **B.** 22Rv1 cells. **C. D.** Intracellular Acetyl-CoA levels are significantly depleted following CBP inhibition. **E.** Lipid droplets were significantly reduced as quantified by BODIPY staining, consistent with impaired fatty acid metabolism downstream of CBP inhibition. **F.** Quantification of lipid droplets by ImageJ. Statistical significance is denoted as \**p* < 0.05, \*\**p* < 0.01, \*\*\**p* < 0.001.

## DISCUSSION

Using a systems-level approach, we leveraged a multimodal, real-world dataset of over 10,000 prostate cancer patients to comprehensively characterize the molecular and clinical evolution from APM-sensitive to -resistant disease and uncover mechanisms of resistance to APM. Integrative analyses of DNA, RNA, and clinical data revealed conserved subnetworks driving resistance and outcomes, including those related to AR signaling, E2F/cell cycle, DNA repair, oxidative phosphorylation, mitochondrial metabolism, and ribosomal function. We identified EP300 and CREB1 as top master regulators of these subnetworks, implicating the CBP/EP300 axis as a central mediator of AR-associated survival signaling in APMR, but AR-altered tumors. Functional studies confirmed that pharmacologic inhibition of CBP/EP300 suppressed prostate cancer cell growth *in vitro* and *in vivo* across multiple AR inhibitor–sensitive and –resistant models with intact AR signaling, but not in models that lacked AR expression. Combined acetylomics and functional studies pinpointed alterations in oxidative phosphorylation and fatty acid metabolism upon exposure to CBP/EP300 inhibition, suggesting a new mechanism of action for pocenbrodib as an inhibitor of oxidative phosphorylation and fatty acid metabolism.

Systems-level analyses have enabled discovery of novel master transcriptional regulators of lethal prostate cancer, such as ONECUT (33), FOXM1 and CENPF (34), GATA2 (35), and MITF (36). The deployment of a systems-level approach in the current work enabled us to illuminate constellations of mechanisms that are coordinately regulated, revealing interactions between networks that may otherwise be concealed. By applying these network inference methods to real-world patient data in the context of both treatment and clinical outcome we were also able to associate these networks with outcomes and, perhaps most importantly, identify key underlying modulators of those outcome-associated mechanisms, leading to identification and clinical translation of a novel CBP/EP300 inhibitor, pocenbrodib, for the treatment of patients with metastatic prostate cancer.

CBP is a key regulator of cellular homeostasis, acting as both a global transcriptional regulator and a nutrient and metabolic sensor. As a histone acetyltransferase, CBP acetylates histone subunits at multiple sites to regulate transcription. Our interrogation of differential acetylation following CBP/EP300 inhibition revealed loss of multiple histone acetylation marks, predominantly in histone 1, 2 and 3 subunits. We also noted loss of acetylation on histone H1X, a linker histone related to maintenance of stemness and neuronal programs (37), and loss of this mark by treatment with CBP/EP300 inhibition may prevent emergence of these aggressive features. In addition to the multiple histone modifications, pocenbrodib also rewired acetylation of multiple non-histone proteins, including a key subset involved in fatty acid metabolism. As a well-established nutrient sensor, CBP/EP300 activity has been shown to regulate hepatic glucose production and lipogenesis (38), glucose uptake by skeletal muscle (39), glucose-dependent browning of white adipose tissue through acetylation of CREBZF (40), and alter pancreatic islet cell metabolism (41). Together, these observations suggest that pocenbrodib may inhibit the capacity of prostate cancer cells to utilize oxidative phosphorylation and fatty acid metabolism as a mechanism of survival in the face of APM by altering acetylation of key ox phos and fatty acid proteins. The key challenge for targeting CBP/EP300 clinically is balancing the profound metabolic inhibition as an anticancer therapy with the metabolic toxicities to normal tissues; trials of pocenbrodib are ongoing in men with mAPMR prostate cancer (NCT06785636) to identify key doses and schedules and combinations to address this.

With clinical translation of multiple novel CBP/EP300 inhibitors underway for the treatment of therapy-resistant prostate cancer, it is critical to understand the underlying mechanisms by which these agents function. A systems-level analysis of multi-scale patient genomics data will help to dissect these mechanisms and identify predictive biomarkers of response to these novel agents.

## METHODS

### Curation of real-world data clinical data

All analyses were performed using de-identified patient data delivered from Tempus, thus making the study exempt from IRB approval. Tempus delivered a cohort of clinical and molecular data during August of 2025 for 12,296 patients that were diagnosed with prostate cancer and received RNA sequencing for at least one tumor tissue sample. The clinical data consisted of structured data curated from individual patient clinical records that were collected as part of the genetic testing ordered on that patient at the time the sample was sent for sequencing. Line of therapy, defined as the first regimen given in the metastatic setting, and regimen information were provided in structured tables which were used for downstream analyses.

### Determining date of castration-resistance

Few patients (6.2%; n = 758) in the delivered cohort have precise date information (MM-YYYY or DD-MM-YYYY precision) for when they were clinically determined to be castration-resistant (CRPC). Thus, we imputed the date of CRPC using the available medication and progression clinical data. The date for which a patient was considered to be CRPC was determined as the first date in the clinical data where a patient experienced a progression event while receiving androgen deprivation therapy (ADT). Progression events were defined as: dictated biochemical recurrence, a dictated progression event (progressive disease), or the identification of a new metastatic site. Eligible progression events were defined as progression events that occurred after the start of the ADT and either before the end of the ADT or in the case of no available end dates, within 3 years of the start of the ADT, under the assumption that patients will be continuing on ADT. Progression events that occurred within 60 days of the start of an ADT were removed to account for potential date shifting (e.g. MM-YYYY precision). The CRPC index was the earliest date for each patient with evidence that the patient was receiving an ADT and had a progression event that matched the above requirements. Using this approach, 2,624 patients (21.3%) were assigned a time of CRPC.

To assign regimens as occurring in either the HSPC or CRPC setting, the regimen was considered to be in the HSPC setting if it was the first regimen in the patient’s record or if the regimen started prior to 60 days before the imputed CRPC date; the regimen was considered to be in the CRPC setting if the regimen was started within 60 days prior or after the imputed CRPC date. To assign samples as collected in either the CRPC or HSPC setting, the sample was assumed to be in the HSPC setting if it was collected prior to the start of the first regimen or prior to the imputed CRPC date and a sample was labeled as CRPC if the sample was collected within 60 days prior or after the imputed CRPC date.

For identification of APM-containing regimens, apalutamide, abiraterone, darolutamide, and enzalutamide were labeled as APM while degarelix, leuprolide, relugolix, goserelin, histrelin, triptorelin, and endocrine therapy were labeled as ADT. Taxanes were defined as docetaxel, palitaxel, or cabazitaxel.

### Outcomes analyses

For outcomes analyses, overall survival was calculated as the time between the reference date (e.g. medication start date) and the time of death. For patients without a death event, the last known follow-up date was used as the event time with a censored flag. When appropriate, left truncation was used to account for immortal time bias. In the case of the overall survival analyses independent of tissue biopsy information, the earliest reception date for a biopsy in the Tempus data was used as the entry date. In the case of the overall survival analyses utilizing expression of individual genes from the tumor tissue, the biopsy date of the tissue was used as the entry date. All outcomes analyses were performed in R version 4.2.1 using the coxph function in the survival library (version 3.3.1).

### Curation of real-world data molecular data

Molecular data (e.g. DNA variant and copy number variant (CNV) calls, normalized RNA-sequencing counts and TPMs when applicable) were delivered for tumor samples (n = 12,781) available from each patient in the cohort. For DNA sequencing, samples were sequenced on either a targeted panel with sequencing data on ∼600 genes (xT, xT.v2, xT.v3, xT.v4) (42) or a more expansive panel (xO, xE, xE.v2). The majority of samples (n = 11,702; 91.6%) were sequenced on xT.v4. Analyses utilizing DNA sequencing information were limited to genes that were measured on xT.v4, where if a gene was not measured on the panel used to sequence the sample, the sample was dropped from the analysis. For RNA sequencing, samples were sequenced on either RS (n = 1,142; 8.9%) or RS.v2 (n = 11,466; 89.7%).

DNA alterations were determined based on DNA mutations and copy number variations (CNVs). DNA mutations were short variants classified as either pathogenic or likely pathogenic based on the Tempus pipeline. DNA copy number variations were labeled as loss if the average length-adjusted copy number across probes covering the gene was <= 0.5 copies or gain if the average length-adjusted copy number across probes covering the gene was > 7.5 copies; all other samples were labeled as wild-type (WT). The RNA-sequencing data consisted of raw gene counts and normalized gene TPMs, which account for differences between the RNA assays. Unless otherwise noted, all outcomes analyses included the DNA or RNA assay to account for biases between overall survival and the assay used. In addition, all differential expression analyses included both assay version (RS vs RS.v2) as well as the tissue site that the biopsy was collected from as co-variates.

### Construction of regulatory networks

Networks, consisting of genes as nodes and directed and undirected edges, termed hybrid networks, were constructed incorporating information derived from weighted gene coexpression analysis (WGCNA), causal mediation (CM) analysis, and Bayesian networks (BNs). While networks were constructed for samples from various treatment and clinical status groups of interest, this manuscript focuses on the two networks (termed the pre-treatment and post-APM networks) built from samples labeled as either pre-ADT/pre-ARSI or post-ARSI, representing early and late-stage disease respectively.

Weighted gene coexpression networks (WGCNs) were constructed using the WGCNA package in R (43). Samples were excluded based on clustering by euclidean distance (using the parDist function from the parallelDist package in R) and clustering using complete linkage. Scale free independence beta soft thresholds were 13.5 for both the pre-ADT/pre-ARSI and post-ARSI WGCNs. Bayesian networks were constructed using RIMBANET (30-32) with 8,000 nodes, where nodes (genes) were selected based on the top varying genes in the individual sets of samples.

### Construction of latent-variable layer

One limitation of PCNs is their reliance on gene expression levels as a surrogate for the regulatory programs the corresponding genes may induce (e.g., the activity levels of transcription factors reflected through their gene expression levels at the time the tumors were biopsied). This assumption can fail when the expression levels of, say, TF or TF coactivators are poorly correlated or even uncorrelated with their actual activity (e.g., when a TF’s activity is regulated post-transcriptionally or post-translationally). To overcome these limitations, we extend the PCN framework by incorporating a latent-variable layer derived from the RNs, to boost key driver analyses in the PCNs, ensuring that the TF-target relationships are grounded in empirical data rather than relying solely on correlations inferred from expression levels. With the resulting hierarchical network, we carried out a key driver analysis to identify key driver genes of each of the FORCE subnetworks.

To construct the latent-variable layer, for each TF or TF coactivator, *T_i_*, we employed a random-walk with restart (RWR) algorithm to generate regulatory influence scores, *s*(*T_i_*, *G_j_*), that reflect the influence of *T_i_* on genes *G_j_* as determined in the RN. This method allows us to quantify both direct and indirect regulatory effects by simulating how a TF’s influence propagates through the network, probabilistically considering all possible paths between *T_i_*and *G_j_* to establish the relationship. The influence scores were then integrated into the PCN key driver analyses using a hierarchical approach, where we first identified the key drivers of each module *M_k_* of interest (for *k* = 1, ... , *L*) using the PCN, and then we identified whether those genes *G_j_* influenced by each *T_i_* in the RNA, were key driver genes in PCN for *M_k_* in the PCN. If such *G_j_* were identified as a key driver gene, we again used the RWR algorithm to compute key driver influence scores across all genes in *M_k_*, resulting in a hierarchically weighted enrichment score across all key drivers of the module *M_k_* of interest:

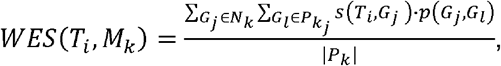

where *p* is the stationary probability score of gene *G_j_*’s influence on *G_l_* in the PCN, *N_k_* is the set of genes *G_j_* in which *T′_i_s* influence is > 0, *P_k_j__* is the set of genes *G_l_* in which gene *G_j_′s* influence is > 0 (*j* = 1, ... ,|*N_k_*|), and *P_k_* is the set of genes in *N_k_* ∪ *P_k_j__* that have both *s* and *p* influence scores > 0. To establish *T_i_* as a key driver in the PCN through this hierarchical approach, we tested whether the set of genes in *P_k_* was enriched for genes in the module *M_k_* using a Fisher Exact Test. Finally, to identify the top-hitting key driver genes across all modules of interest, we rank-ordered all key driver genes identified directly through the PCN or through the hierarchical network approach using a composite score constructed from -log10 of the p-values for the individual Fisher Exact Tests applied to each module.

### Cell lines

LNCaP cells were obtained from ATCC via the Duke University Cell Culture Facility. LNCaP cells were cultured in RPMI media supplemented with 10% fetal bovine serum (Sigma) and 1% penicillin/streptomycin (Life Technologies). LNCaP95 cells were a gift of Dr. Scott Dehm (University of Minnesota). Both CS2 and LNCaP95 cells are androgen-independent lines derived from a parental LNCaP line and grown in charcoal stripped serum. They are grown in RPMI supplemented with 10% charcoal stripped fetal bovine serum and 1% penicillin/streptomycin. Enzalutamide resistant (Enza-R) cell lines were grown in steadily increasing doses of enzalutamide until a maximum concentration of 50 μM was reached. A sister line of each was carried in DMSO at equal passage number to their Enza-R counterpart. Each cell line was authenticated both before and after chronic exposure to drug or vehicle. Cells expressing Snail (Addgene plasmid #18798) were created as previously described (44).

### Cell growth and viability assays

Cells were grown in the presence of DMSO or 50 μM enzalutamide for at least one week prior to the start of any experiment. Cells were then counted using the Countess II (Life Technologies) and plated in a 96 well plate at a concentration of 2500 cells/well in 100 μL of media. 24 hours after the initial plating, cells were dosed with a 10-point dose curve at a 1:2 dilution, starting at 50 μM CBP/EP300 inhibitor and ending with a DMSO control well. Cells were dosed with 100 μL at 2X concentration to reach the final desired concentration. Cell growth and confluence was monitored using the IncuCyte live cell analysis system (Essen Biosciences) and each experiment was considered to have reached its endpoint when the DMSO control wells reach ∼90% confluency. When the endpoint was reached, CellTiter-Glo (Promega) was added to the desired wells and imaged on the microplate reader (Molecular Devices) using the accompanying software.

### Quantitative RT-PCR

Cells were treated for 24 hours with 1 μM CBP/EP300 inhibitor and then RNA was extracted using the Quick-RNA Miniprep kit (Zymo Research). Total RNA was then reverse transcribed using the High-Capacity cDNA Reverse Transcription Kit (Life Technologies). A five-fold dilution of the reactions was created and used in a quantitative (q)PCR reaction with Sybr Master Mix. The reaction was run using the Vii7 real time-QPCR detection system (Applied Biosystems). GAPDH mRNA levels were used to normalize all other mRNA levels and presented as “relative expression levels”. The complete list of primer sequences is included in the supplementary text.

### Immunoblot analyses

Cells were treated for 24 hours with 1 μM CBP/EP300 inhibitor and lysed using either the Histone Extraction Kit (Abcam) or RIPA buffer with protease and phosphatase inhibitor (Thermo Fisher). Samples were then treated in accordance with NuPage protocols, using DTT as the reducing agent. Samples were then loaded into a 4-12% Bis-Tris gel with MOPS running buffer (Thermo Fisher) and samples were run for 40 minutes at 200 volts. Once the run was completed, the gels were transferred onto nitrocellulose and blocked in StartingBlock (Thermo), incubated with antibodies, and imaged using the Odyssey-FC Imager (LI-COR).

### Colony forming and spheroid assays

A total of 2,500 cells were plated in each well of a 6-well plate and incubated overnight. The following day, the media was replaced with either media containing 1 μM CBP/EP300 inhibitor or the equivalent volume of DMSO. Cells were incubated for 14 days before the media was removed. Cells were then washed with PBS and stained with Crystal Violet. For spheroid assays, 2,500 cells were plated into each well of a 384 ultra-low attachment spheroid plate (Corning) and centrifuged at >500g for 5 minutes to aid in spheroid formation. Drug was added day 1 of the experiment, prior to centrifugation. Cells were then imaged daily using the IncuCyte live cell analysis system (Essen Biosciences). Images were exported and analyzed in ImageJ to determine the area of the spheroid surface.

### LC-MS/MS Sample Preparation

Cells were treated for 24 h with DMSO (vehicle) or 1 μM CBP/EP300 inhibitor (FT-6876). They were then scraped in 8 M urea/50 mM ammonium bicarbonate to a final protein concentration of 10 mg/mL in a total volume of 1 mL and sonicated with a probe three times for 5 s each at 30% energy. Lysates were centrifuged at 12,000 × g for 5 min at 4 °C. Protein concentrations of the supernatants were determined in duplicate by Bradford assay.

For acetylomics, 12 mg of protein from each sample was used; for expression proteomics, 50 μg was used. Samples were normalized with 8 M urea in 50 mM ammonium bicarbonate, reduced with 10 mM dithiothreitol for 45 min at 32 °C, and alkylated with 25 mM iodoacetamide for 30 min at room temperature. Samples were then diluted to 1.6 M urea with 50 mM ammonium bicarbonate and digested with trypsin at a 1:50 enzyme-to-protein ratio for 18 h at 32 °C. Digests were acidified with TFA, cleaned up by C18 SPE (Sep-Pak, 500 mg bed), and lyophilized to dryness. For acetylomics, acetylated peptides were enriched by immunoaffinity purification using the PTMScan kit 13416 (Cell Signaling Technology), following the manufacturer’s recommended IP, washing and elution protocol. Eluates were lyophilized, resuspended in 2% acetonitrile/0.1% TFA, cleaned up with stage tips, and lyophilized again.

Before LC-MS/MS, samples were resuspended in 1% TFA/2% acetonitrile containing yeast alcohol dehydrogenase (ADH_YEAST) as an internal QC control. Acetyl-enriched samples were resuspended in 8 μL containing 12.5 or 25 fmol/μL ADH, and expression samples in 100 μL containing 25 fmol/μL ADH. For each experiment, a study pool QC (SPQC) sample was made by combining equal aliquots of every sample: 2 μL per acetylomics sample and 10 μL per expression sample. This pool was analyzed periodically throughout acquisition.

### LC-MS/MS Data Collection and Data Analysis

Quantitative LC-MS/MS was performed on 4.5 μL of each acetyl-enriched sample and 2 μL of each expression sample on a Thermo Orbitrap Fusion Lumos high-resolution accurate-mass tandem mass spectrometer. Samples were first trapped on a Symmetry C18 20 mm × 180 μm trapping column at 5 μL/min in 99.9/0.1 v/v water/acetonitrile. Analytical separation used a 1.8 μm Acquity HSS T3 C18 75 μm × 250 mm column (Waters Corp.) with a 90-min linear gradient of 5–30% acetonitrile with 0.1% formic acid, at 400 nL/min and 55 °C. Data were collected in data-dependent acquisition (DDA) mode. Full MS scans were acquired at r = 120,000 (at m/z 200) from m/z 375–1500 with an AGC target of 2e5 ions. MS/MS scans were acquired in the ion trap at the Rapid scan rate, with an AGC target of 5e3 ions and a maximum injection time of 100 ms. The total cycle time for MS and MS/MS scans was 2s, and a 20s dynamic exclusion was used to increase depth of coverage.

Each experiment comprised 15 UPLC-MS/MS analyses including 3 SPQC injections, one after every 6 samples across the sample set. Data were imported into Proteome Discoverer (v2.4). All runs were aligned by the accurate mass and retention time of detected ions (“features”) that contained MS/MS spectra, using the Minora Feature Detector algorithm. Fragment ion spectra were produced and searched with Mascot Distiller and Mascot Server (v2.5, Matrix Science). Searches were run against the SwissProt *H. sapiens* database plus an equal number of reversed-sequence decoys for false discovery rate (FDR) determination. Both searches used a fixed modification of carbamidomethylation on Cys and variable oxidation on Met. The acetylomics search added variable acetylation on Lys and protein N-termini. The expression search added variable deamidation on Asn and Gln. Peptides were annotated at a maximum 1% peptide spectral match (PSM) FDR (**Supplemental Tables 2 and 3**). For expression data, the Peptide Validator and Protein FDR Validator nodes were also used to annotate the data at a maximum 1% protein FDR.

Relative peptide abundance was calculated from the area under the curve (AUC) of the selected ion chromatograms of aligned features across all runs. Data were normalized by sample loading and by trimmed mean (excluding the top and bottom 10%), and missing values were then imputed. If fewer than half the values in a treatment group were missing, each was imputed with an intensity drawn from a normal distribution of measured values in the same intensity range (20 bins). If more than half the values were missing for a peptide in a group and its intensity was > 5e^6^, the peptide was considered misaligned and its measured intensity was set to 0. All remaining missing values were imputed with the lowest 5% of all detected values.

## Data Availability

The de-identified Tempus data used in this study were collected in a real-world healthcare setting and are subject to controlled access for privacy and proprietary reasons; access may be requested from Tempus AI, Inc. and is subject to a data use agreement. Derived data supporting the findings of this study are provided within the manuscript and its supplementary materials. All other data produced in the present study are available upon reasonable request to the authors.

## SUPPLEMENTARY FIGURE LEGENDS

**Supplemental Figure 1.**
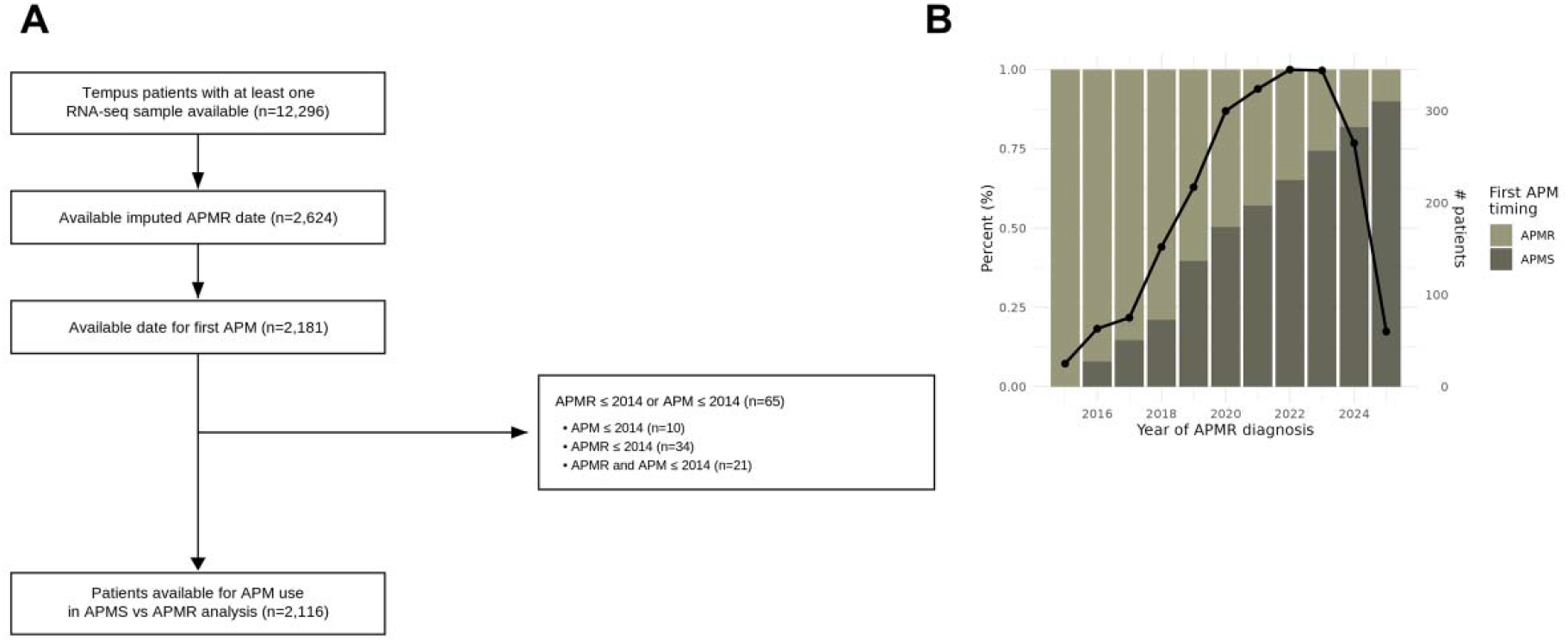
APM use has shifted from the mAPMR setting to the mAPMS setting over time in real-world practice. (A) CONSORT diagram depicting selection of 2,116 patients from the Tempus data with available imputed APMR dates and a record of a first APM that were used to characterize changes in temporal use of APMs across disease settings. (B) After binning patients into either the APM in the APMS or APMR setting groups, the proportion of patients receiving an APM for the first time in the APMS setting has increased substantially from 2016 onward.

**Supplemental Figure 2.**
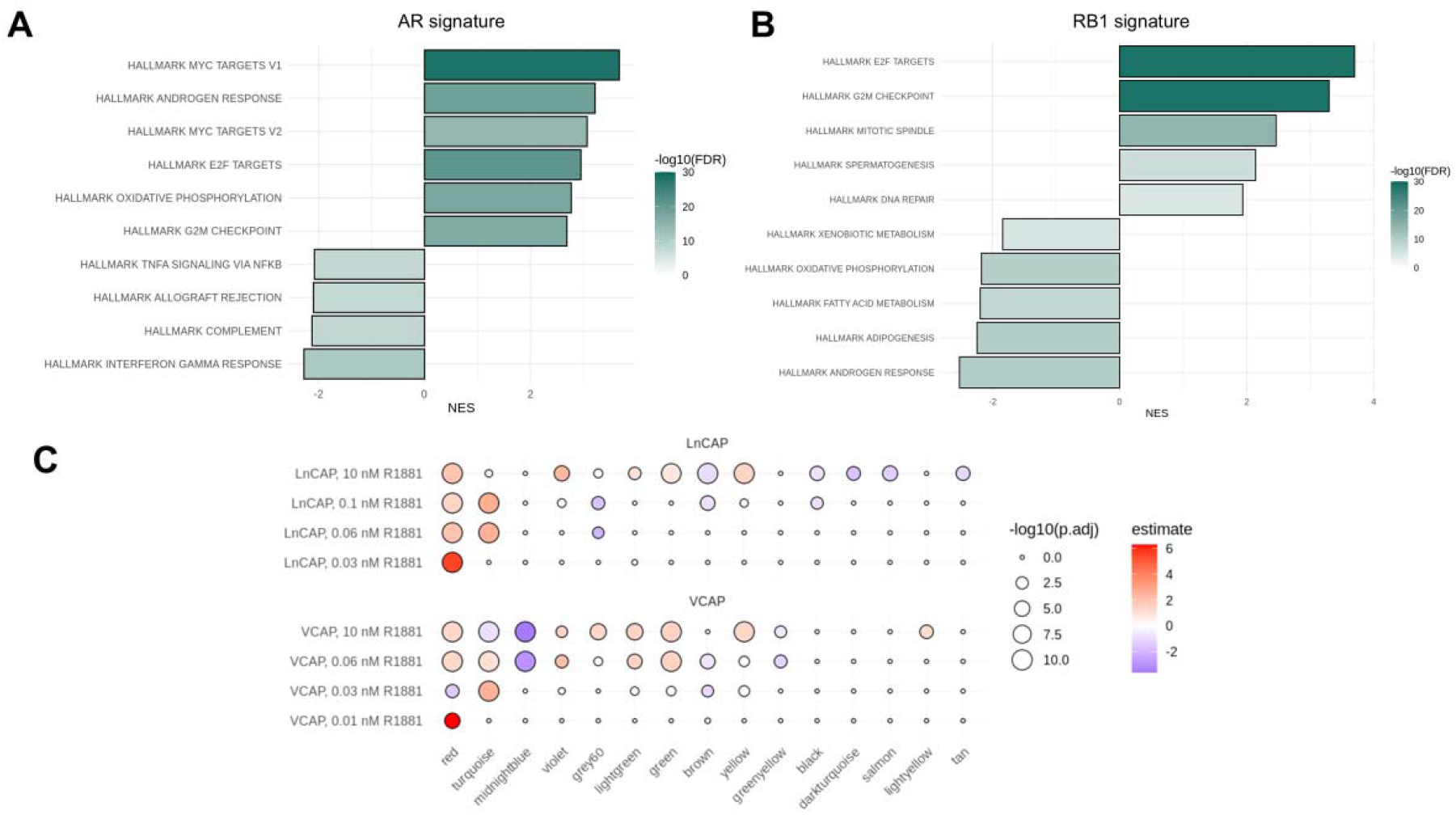
Gene set enrichment analysis (GSEA) of differentially expressed genes in the AR and RB1 signatures using Hallmark pathways. (**A**) AR-altered tumors are enriched for MYC targets, Hallmark Androgen Response, E2F targets, and oxidative phosphorylation, consistent with AR amplification driving coordinated upregulation of proliferative and metabolic transcriptional programs. (**B**) RB1-altered tumors are enriched for E2F targets, G2M checkpoint, mitotic spindle, and spermatogenesis gene sets, consistent with RB1 loss-driven de-repression of cell cycle and activation of neuroendocrine-associated transcriptional programs. (**C**) Validation of AR signature-associated subnetworks using publicly available RNA-seq data from LNCaP and VCaP cell lines treated with varying concentrations of the synthetic androgen R1881.

**Supplemental Figure 3.**
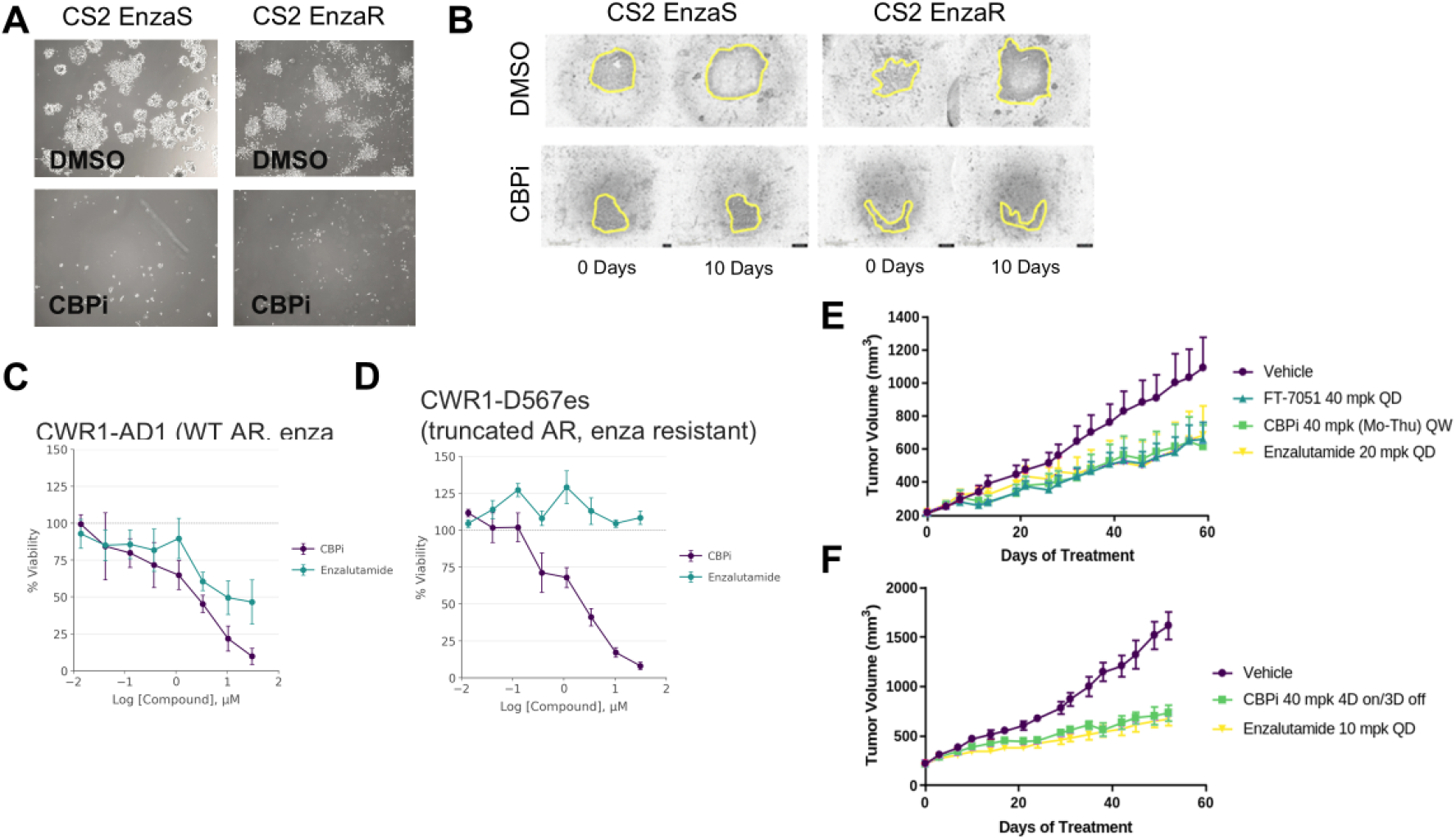
Evaluation of the efficacy of CBP/EP300 inhibition in APM-resistant prostate cancer. (**A**) Colony forming assays for LNCaP-CS2 enzalutamide-sensitive and –resistant lines treated with vehicle (DMSO) or tool compound, FT-6876 (CBPi). (**B**) Spheroid assays in LNCaP-CS2 enzalutamide-sensitive and –resistant lines treated with vehicle (DMSO) or FT-6876 (CBPi). (**C**) Inhibition curves in CWR1-AD1 cells with wild type AR treated with enzalutamide or tool compound, FT-6876. (**D**) Enzalutamide-resistant CWR1-567es cells with truncated AR treated with enzalutamide or tool compound FT-6876. (**E**) *In vivo* efficacy of CBPi, FT-6876, in enzalutamide-sensitive PDXs, PDX- and (**F**) PDX-

## REFERENCES

1. de Bono J, Mateo J, Fizazi K, Saad F, Shore N, Sandhu S, Chi KN, Sartor O, Agarwal N, Olmos D, Thiery-Vuillemin A, Twardowski P, Mehra N, Goessl C, Kang J, Burgents J, Wu W, Kohlmann A, Adelman CA, Hussain M. Olaparib for Metastatic Castration-Resistant Prostate Cancer. N Engl J Med. 2020;382(22):2091–102. Epub 2020/04/29. doi: 10.1056/NEJMoa1911440. PubMed PMID: 32343890.https://www.ncbi.nlm.nih.gov/pubmed/32343890

2. Fizazi K, Azad AA, Matsubara N, Carles J, Fay AP, De Giorgi U, Joung JY, Fong PCC, Voog E, Jones RJ, Shore ND, Dunshee C, Zschabitz S, Oldenburg J, Ye D, Lin X, Healy CG, Di Santo N, Laird AD, Zohren F, Agarwal N. First-line talazoparib with enzalutamide in HRR-deficient metastatic castration-resistant prostate cancer: the phase 3 TALAPRO-2 trial. Nat Med. 2024;30(1):257–64. Epub 2023/12/05. doi: 10.1038/s41591-023-02704-x. PubMed PMID: 38049622; PMCID: PMC10803259 https://www.ncbi.nlm.nih.gov/pubmed/38049622

3. Saad F, Clarke NW, Oya M, Shore N, Procopio G, Guedes JD, Arslan C, Mehra N, Parnis F, Brown E, Schlurmann F, Joung JY, Sugimoto M, Sartor O, Liu YZ, Poehlein C, Barker L, Del Rosario PM, Armstrong AJ. Olaparib plus abiraterone versus placebo plus abiraterone in metastatic castration-resistant prostate cancer (PROpel): final prespecified overall survival results of a randomised, double-blind, phase 3 trial. Lancet Oncol. 2023;24(10):1094–108. Epub 2023/09/16. doi: 10.1016/S1470-2045(23)00382-0. PubMed PMID: 37714168.https://www.ncbi.nlm.nih.gov/pubmed/37714168

4. Spratt DE, Yousefi K, Deheshi S, Ross AE, Den RB, Schaeffer EM, Trock BJ, Zhang J, Glass AG, Dicker AP, Abdollah F, Zhao SG, Lam LLC, du Plessis M, Choeurng V, Haddad Z, Buerki C, Davicioni E, Weinmann S, Freedland SJ, Klein EA, Karnes RJ, Feng FY. Individual Patient-Level Meta-Analysis of the Performance of the Decipher Genomic Classifier in High-Risk Men After Prostatectomy to Predict Development of Metastatic Disease. J Clin Oncol. 2017;35(18):1991–8. Epub 2017/03/31. doi: 10.1200/JCO.2016.70.2811. PubMed PMID: 28358655; PMCID: PMC6530581.https://www.ncbi.nlm.nih.gov/pubmed/28358655

5. Cronin KA, Scott S, Firth AU, Sung H, Henley SJ, Sherman RL, Siegel RL, Anderson RN, Kohler BA, Benard VB, Negoita S, Wiggins C, Cance WG, Jemal A. Annual report to the nation on the status of cancer, part 1: National cancer statistics. Cancer. 2022;128(24):4251–84. Epub 20221027. doi: 10.1002/cncr.34479. PubMed PMID: 36301149; PMCID: PMC10092838.https://www.ncbi.nlm.nih.gov/pubmed/36301149

6. Armstrong AJ, Morris MJ, Abida W, Aggarwal RR, Antonarakis ES, Attard G, Beltran H, Bryce A, Carducci MA, Cheng HH, Chen DL, Chi KN, Childs DS, Dahut W, Emmett L, Fizazi K, Gafita A, George DJ, Hermann K, Hofman MS, Hope T, Hussain M, Kelly WK, Kessler E, Kuo PH, Lang J, Liu G, Marshall CH, Morgans AK, McKay RR, Nanus D, Nelson P, Paller C, Reichert ZR, Ryan CJ, Sartor AO, Schoder H, Schwartz LH, Sharifi N, Stadler WM, Stein M, Sternberg CN, Szmulewitz RZ, Tagawa ST, Sokolova AO, Wyatt AW, Yamoah K, Yu EY, Halabi S, Scher HI, Group PW. Trial Design and Objectives for Patients With Prostate Cancer: Recommendations From the Prostate Cancer Working Group 4. J Clin Oncol. 2026;44(13):1249–65. Epub 2026/02/26. doi: 10.1200/JCO-25-02834. PubMed PMID: 41744290; PMCID: PMC13132095 https://www.ncbi.nlm.nih.gov/pubmed/41744290

7. Armstrong AJ, Azad AA, Iguchi T, Szmulewitz RZ, Petrylak DP, Holzbeierlein J, Villers A, Alcaraz A, Alekseev B, Shore ND, Gomez-Veiga F, Rosbrook B, Zohren F, Yamada S, Haas GP, Stenzl A. Improved Survival With Enzalutamide in Patients With Metastatic Hormone-Sensitive Prostate Cancer. J Clin Oncol. 2022;40(15):1616–22. Epub 2022/04/15. doi: 10.1200/JCO.22.00193. PubMed PMID: 35420921; PMCID: PMC9113211.https://www.ncbi.nlm.nih.gov/pubmed/35420921

8. Chi KN, Chowdhury S, Bjartell A, Chung BH, Pereira de Santana Gomes AJ, Given R, Juarez A, Merseburger AS, Ozguroglu M, Uemura H, Ye D, Brookman-May S, Mundle SD, McCarthy SA, Larsen JS, Sun W, Bevans KB, Zhang K, Bandyopadhyay N, Agarwal N. Apalutamide in Patients With Metastatic Castration-Sensitive Prostate Cancer: Final Survival Analysis of the Randomized, Double-Blind, Phase III TITAN Study. J Clin Oncol. 2021;39(20):2294–303. Epub 2021/04/30. doi: 10.1200/JCO.20.03488. PubMed PMID: 33914595.https://www.ncbi.nlm.nih.gov/pubmed/33914595

9. Chi KN, Agarwal N, Bjartell A, Chung BH, Pereira de Santana Gomes AJ, Given R, Juarez Soto A, Merseburger AS, Ozguroglu M, Uemura H, Ye D, Deprince K, Naini V, Li J, Cheng S, Yu MK, Zhang K, Larsen JS, McCarthy S, Chowdhury S, Investigators T. Apalutamide for Metastatic, Castration-Sensitive Prostate Cancer. N Engl J Med. 2019;381(1):13–24. Epub 2019/06/01. doi: 10.1056/NEJMoa1903307. PubMed PMID: 31150574.https://www.ncbi.nlm.nih.gov/pubmed/31150574

10. Fizazi K, Tran N, Fein L, Matsubara N, Rodriguez-Antolin A, Alekseev BY, Ozguroglu M, Ye D, Feyerabend S, Protheroe A, De Porre P, Kheoh T, Park YC, Todd MB, Chi KN, Investigators L. Abiraterone plus Prednisone in Metastatic, Castration-Sensitive Prostate Cancer. N Engl J Med. 2017;377(4):352–60. Epub 2017/06/06. doi: 10.1056/NEJMoa1704174. PubMed PMID: 28578607.https://www.ncbi.nlm.nih.gov/pubmed/28578607

11. Clarke NW, Ali A, Ingleby FC, Hoyle A, Amos CL, Attard G, Brawley CD, Calvert J, Chowdhury S, Cook A, Cross W, Dearnaley DP, Douis H, Gilbert D, Gillessen S, Jones RJ, Langley RE, MacNair A, Malik Z, Mason MD, Matheson D, Millman R, Parker CC, Ritchie AWS, Rush H, Russell JM, Brown J, Beesley S, Birtle A, Capaldi L, Gale J, Gibbs S, Lydon A, Nikapota A, Omlin A, O’Sullivan JM, Parikh O, Protheroe A, Rudman S, Srihari NN, Simms M, Tanguay JS, Tolan S, Wagstaff J, Wallace J, Wylie J, Zarkar A, Sydes MR, Parmar MKB, James ND. Addition of docetaxel to hormonal therapy in low- and high-burden metastatic hormone sensitive prostate cancer: long-term survival results from the STAMPEDE trial. Ann Oncol. 2019;30(12):1992–2003. Epub 2019/09/29. doi: 10.1093/annonc/mdz396. PubMed PMID: 31560068; PMCID: PMC6938598.https://www.ncbi.nlm.nih.gov/pubmed/31560068

12. Parker CC, James ND, Brawley CD, Clarke NW, Hoyle AP, Ali A, Ritchie AWS, Attard G, Chowdhury S, Cross W, Dearnaley DP, Gillessen S, Gilson C, Jones RJ, Langley RE, Malik ZI, Mason MD, Matheson D, Millman R, Russell JM, Thalmann GN, Amos CL, Alonzi R, Bahl A, Birtle A, Din O, Douis H, Eswar C, Gale J, Gannon MR, Jonnada S, Khaksar S, Lester JF, O’Sullivan JM, Parikh OA, Pedley ID, Pudney DM, Sheehan DJ, Srihari NN, Tran ATH, Parmar MKB, Sydes MR, Systemic Therapy for Advanced or Metastatic Prostate cancer: Evaluation of Drug Efficacy i. Radiotherapy to the primary tumour for newly diagnosed, metastatic prostate cancer (STAMPEDE): a randomised controlled phase 3 trial. Lancet. 2018;392(10162):2353-66. Epub 2018/10/26. doi: 10.1016/S0140-6736(18)32486-3. PubMed PMID: 30355464; PMCID: PMC6269599.https://www.ncbi.nlm.nih.gov/pubmed/30355464

13. James ND, de Bono JS, Spears MR, Clarke NW, Mason MD, Dearnaley DP, Ritchie AWS, Amos CL, Gilson C, Jones RJ, Matheson D, Millman R, Attard G, Chowdhury S, Cross WR, Gillessen S, Parker CC, Russell JM, Berthold DR, Brawley C, Adab F, Aung S, Birtle AJ, Bowen J, Brock S, Chakraborti P, Ferguson C, Gale J, Gray E, Hingorani M, Hoskin PJ, Lester JF, Malik ZI, McKinna F, McPhail N, Money-Kyrle J, O’Sullivan J, Parikh O, Protheroe A, Robinson A, Srihari NN, Thomas C, Wagstaff J, Wylie J, Zarkar A, Parmar MKB, Sydes MR, Investigators S. Abiraterone for Prostate Cancer Not Previously Treated with Hormone Therapy. N Engl J Med. 2017;377(4):338–51. Epub 2017/06/06. doi: 10.1056/NEJMoa1702900. PubMed PMID: 28578639; PMCID: PMC5533216.https://www.ncbi.nlm.nih.gov/pubmed/28578639

14. Caligiuri M, al. e. Abstract 3079: FT-6876, a potent and selective inhibitor of CBP/p300 with antitumor activity in AR-positive breast cancer. Cancer Research 2020. p. 3079.

15. Fizazi K, Tran N, Fein L, Matsubara N, Rodriguez-Antolin A, Alekseev BY, Ozguroglu M, Ye D, Feyerabend S, Protheroe A, Sulur G, Luna Y, Li S, Mundle S, Chi KN. Abiraterone acetate plus prednisone in patients with newly diagnosed high-risk metastatic castration-sensitive prostate cancer (LATITUDE): final overall survival analysis of a randomised, double-blind, phase 3 trial. Lancet Oncol. 2019;20(5):686–700. Epub 2019/04/17. doi: 10.1016/S1470-2045(19)30082-8. PubMed PMID: 30987939.https://www.ncbi.nlm.nih.gov/pubmed/30987939

16. Armstrong AJ, Szmulewitz RZ, Petrylak DP, Holzbeierlein J, Villers A, Azad A, Alcaraz A, Alekseev B, Iguchi T, Shore ND, Rosbrook B, Sugg J, Baron B, Chen L, Stenzl A. ARCHES: A Randomized, Phase III Study of Androgen Deprivation Therapy With Enzalutamide or Placebo in Men With Metastatic Hormone-Sensitive Prostate Cancer. J Clin Oncol. 2019;37(32):2974–86. Epub 2019/07/23. doi: 10.1200/JCO.19.00799. PubMed PMID: 31329516; PMCID: PMC6839905.https://www.ncbi.nlm.nih.gov/pubmed/31329516

17. Hussain M, Tombal B, Saad F, Fizazi K, Sternberg CN, Crawford ED, Shore N, Kopyltsov E, Kalebasty AR, Bogemann M, Ye D, Cruz F, Suzuki H, Kapur S, Srinivasan S, Verholen F, Kuss I, Joensuu H, Smith MR. Darolutamide Plus Androgen-Deprivation Therapy and Docetaxel in Metastatic Hormone-Sensitive Prostate Cancer by Disease Volume and Risk Subgroups in the Phase III ARASENS Trial. J Clin Oncol. 2023;41(20):3595–607. Epub 2023/02/17. doi: 10.1200/JCO.23.00041. PubMed PMID: 36795843.https://www.ncbi.nlm.nih.gov/pubmed/36795843

18. Saad F, Vjaters E, Shore N, Olmos D, Xing N, Pereira de Santana Gomes AJ, Cesar de Andrade Mota A, Salman P, Jievaltas M, Ulys A, Jakubovskis M, Kopyltsov E, Han W, Nevalaita L, Testa I, Le Berre MA, Kuss I, Haresh KP, Investigators AS. Darolutamide in Combination With Androgen-Deprivation Therapy in Patients With Metastatic Hormone-Sensitive Prostate Cancer From the Phase III ARANOTE Trial. J Clin Oncol. 2024;42(36):4271–81. Epub 2024/09/16. doi: 10.1200/JCO-24-01798. PubMed PMID: 39279580; PMCID: PMC11654448 https://www.ncbi.nlm.nih.gov/pubmed/39279580

19. Abida W, Cyrta J, Heller G, Prandi D, Armenia J, Coleman I, Cieslik M, Benelli M, Robinson D, Van Allen EM, Sboner A, Fedrizzi T, Mosquera JM, Robinson BD, De Sarkar N, Kunju LP, Tomlins S, Wu YM, Nava Rodrigues D, Loda M, Gopalan A, Reuter VE, Pritchard CC, Mateo J, Bianchini D, Miranda S, Carreira S, Rescigno P, Filipenko J, Vinson J, Montgomery RB, Beltran H, Heath EI, Scher HI, Kantoff PW, Taplin ME, Schultz N, deBono JS, Demichelis F, Nelson PS, Rubin MA, Chinnaiyan AM, Sawyers CL. Genomic correlates of clinical outcome in advanced prostate cancer. Proc Natl Acad Sci U S A. 2019;116(23):11428–36. Epub 2019/05/08. doi: 10.1073/pnas.1902651116. PubMed PMID: 31061129; PMCID: PMC6561293.https://www.ncbi.nlm.nih.gov/pubmed/31061129

20. Abida W, Cheng ML, Armenia J, Middha S, Autio KA, Vargas HA, Rathkopf D, Morris MJ, Danila DC, Slovin SF, Carbone E, Barnett ES, Hullings M, Hechtman JF, Zehir A, Shia J, Jonsson P, Stadler ZK, Srinivasan P, Laudone VP, Reuter V, Wolchok JD, Socci ND, Taylor BS, Berger MF, Kantoff PW, Sawyers CL, Schultz N, Solit DB, Gopalan A, Scher HI. Analysis of the Prevalence of Microsatellite Instability in Prostate Cancer and Response to Immune Checkpoint Blockade. JAMA Oncol. 2019;5(4):471–8. Epub 2018/12/28. doi: 10.1001/jamaoncol.2018.5801. PubMed PMID: 30589920; PMCID: PMC6459218.https://www.ncbi.nlm.nih.gov/pubmed/30589920

21. Mu P, Zhang Z, Benelli M, Karthaus WR, Hoover E, Chen CC, Wongvipat J, Ku SY, Gao D, Cao Z, Shah N, Adams EJ, Abida W, Watson PA, Prandi D, Huang CH, de Stanchina E, Lowe SW, Ellis L, Beltran H, Rubin MA, Goodrich DW, Demichelis F, Sawyers CL. SOX2 promotes lineage plasticity and antiandrogen resistance in TP53- and RB1-deficient prostate cancer. Science. 2017;355(6320):84–8. Epub 2017/01/07. doi: 10.1126/science.aah4307. PubMed PMID: 28059768; PMCID: PMC5247742.https://www.ncbi.nlm.nih.gov/pubmed/28059768

22. Ku SY, Rosario S, Wang Y, Mu P, Seshadri M, Goodrich ZW, Goodrich MM, Labbe DP, Gomez EC, Wang J, Long HW, Xu B, Brown M, Loda M, Sawyers CL, Ellis L, Goodrich DW. Rb1 and Trp53 cooperate to suppress prostate cancer lineage plasticity, metastasis, and antiandrogen resistance. Science. 2017;355(6320):78–83. Epub 2017/01/07. doi: 10.1126/science.aah4199. PubMed PMID: 28059767; PMCID: PMC5367887.https://www.ncbi.nlm.nih.gov/pubmed/28059767

23. Beltran H, Prandi D, Mosquera JM, Benelli M, Puca L, Cyrta J, Marotz C, Giannopoulou E, Chakravarthi BV, Varambally S, Tomlins SA, Nanus DM, Tagawa ST, Van Allen EM, Elemento O, Sboner A, Garraway LA, Rubin MA, Demichelis F. Divergent clonal evolution of castration-resistant neuroendocrine prostate cancer. Nat Med. 2016;22(3):298–305. Epub 2016/02/09. doi: 10.1038/nm.4045. PubMed PMID: 26855148; PMCID: PMC4777652.https://www.ncbi.nlm.nih.gov/pubmed/26855148

24. Aggarwal R, Huang J, Alumkal JJ, Zhang L, Feng FY, Thomas GV, Weinstein AS, Friedl V, Zhang C, Witte ON, Lloyd P, Gleave M, Evans CP, Youngren J, Beer TM, Rettig M, Wong CK, True L, Foye A, Playdle D, Ryan CJ, Lara P, Chi KN, Uzunangelov V, Sokolov A, Newton Y, Beltran H, Demichelis F, Rubin MA, Stuart JM, Small EJ. Clinical and Genomic Characterization of Treatment-Emergent Small-Cell Neuroendocrine Prostate Cancer: A Multi-institutional Prospective Study. J Clin Oncol. 2018;36(24):2492–503. Epub 2018/07/10. doi: 10.1200/JCO.2017.77.6880. PubMed PMID: 29985747; PMCID: PMC6366813.https://www.ncbi.nlm.nih.gov/pubmed/29985747

25. Safi R, Wardell SE, Watkinson P, Qin X, Lee M, Park S, Krebs T, Dolan EL, Blattler A, Tsuji T, Nayak S, Khater M, Fontanillo C, Newlin MA, Kirkland ML, Xie Y, Long H, Fink EC, Fanning SW, Runyon S, Brown M, Xu S, Owzar K, Norris JD, McDonnell DP. Androgen receptor monomers and dimers regulate opposing biological processes in prostate cancer cells. Nat Commun. 2024;15(1):7675. Epub 2024/09/04. doi: 10.1038/s41467-024-52032-y. PubMed PMID: 39227594; PMCID: PMC11371910.https://www.ncbi.nlm.nih.gov/pubmed/39227594

26. Ayers KL, Ma M, Debussche G, Corrigan D, McCafferty J, Lee K, Newman S, Zhou X, Hirsch FR, Mack PC, Liu JJ, Schadt EE, Chen R, Li SD. A composite biomarker of neutrophil-lymphocyte ratio and hemoglobin level correlates with clinical response to PD-1 and PD-L1 inhibitors in advanced non-small cell lung cancers. BMC Cancer. 2021;21(1):441. Epub 20210421. doi: 10.1186/s12885-021-08194-9. PubMed PMID: 33882890; PMCID: PMC8059160.https://www.ncbi.nlm.nih.gov/pubmed/33882890

27. Ma M, Wang C, Glicksberg BS, Schadt EE, Li SD, Chen R. Identify Cancer Driver Genes through Shared Mendelian Disease Pathogenic Variants and Cancer Somatic Mutations. Pac Symp Biocomput. 2017;22:473–84. doi: 10.1142/9789813207813_0044. PubMed PMID: 27896999.https://www.ncbi.nlm.nih.gov/pubmed/27896999

28. Uzilov AV, Ding W, Fink MY, Antipin Y, Brohl AS, Davis C, Lau CY, Pandya C, Shah H, Kasai Y, Powell J, Micchelli M, Castellanos R, Zhang Z, Linderman M, Kinoshita Y, Zweig M, Raustad K, Cheung K, Castillo D, Wooten M, Bourzgui I, Newman LC, Deikus G, Mathew B, Zhu J, Glicksberg BS, Moe AS, Liao J, Edelmann L, Dudley JT, Maki RG, Kasarskis A, Holcombe RF, Mahajan M, Hao K, Reva B, Longtine J, Starcevic D, Sebra R, Donovan MJ, Li S, Schadt EE, Chen R. Development and clinical application of an integrative genomic approach to personalized cancer therapy. Genome Med. 2016;8(1):62. Epub 20160601. doi: 10.1186/s13073-016-0313-0. PubMed PMID: 27245685; PMCID: PMC4888213.https://www.ncbi.nlm.nih.gov/pubmed/27245685

29. Wang L, Sfakianos JP, Beaumont KG, Akturk G, Horowitz A, Sebra RP, Farkas AM, Gnjatic S, Hake A, Izadmehr S, Wiklund P, Oh WK, Szabo PM, Wind-Rotolo M, Unsal-Kacmaz K, Yao X, Schadt E, Sharma P, Bhardwaj N, Zhu J, Galsky MD. Myeloid Cell-associated Resistance to PD-1/PD-L1 Blockade in Urothelial Cancer Revealed Through Bulk and Single-cell RNA Sequencing. Clin Cancer Res. 2021;27(15):4287–300. Epub 20210409. doi: 10.1158/1078-0432.CCR-20-4574. PubMed PMID: 33837006; PMCID: PMC8338756.https://www.ncbi.nlm.nih.gov/pubmed/33837006

30. Zhu J, Sova P, Xu Q, Dombek KM, Xu EY, Vu H, Tu Z, Brem RB, Bumgarner RE, Schadt EE. Stitching together multiple data dimensions reveals interacting metabolomic and transcriptomic networks that modulate cell regulation. PLoS Biol. 2012;10(4):e1001301. Epub 20120403. doi: 10.1371/journal.pbio.1001301. PubMed PMID: 22509135; PMCID: PMC3317911.https://www.ncbi.nlm.nih.gov/pubmed/22509135

31. Zhu J, Wiener MC, Zhang C, Fridman A, Minch E, Lum PY, Sachs JR, Schadt EE. Increasing the power to detect causal associations by combining genotypic and expression data in segregating populations. PLoS Comput Biol. 2007;3(4):e69. Epub 20070227. doi: 10.1371/journal.pcbi.0030069. PubMed PMID: 17432931; PMCID: PMC1851982.https://www.ncbi.nlm.nih.gov/pubmed/17432931

32. Zhu J, Zhang B, Smith EN, Drees B, Brem RB, Kruglyak L, Bumgarner RE, Schadt EE. Integrating large-scale functional genomic data to dissect the complexity of yeast regulatory networks. Nat Genet. 2008;40(7):854–61. Epub 20080615. doi: 10.1038/ng.167. PubMed PMID: 18552845; PMCID: PMC2573859.https://www.ncbi.nlm.nih.gov/pubmed/18552845

33. Rotinen M, You S, Yang J, Coetzee SG, Reis-Sobreiro M, Huang WC, Huang F, Pan X, Yanez A, Hazelett DJ, Chu CY, Steadman K, Morrissey CM, Nelson PS, Corey E, Chung LWK, Freedland SJ, Di Vizio D, Garraway IP, Murali R, Knudsen BS, Freeman MR. ONECUT2 is a targetable master regulator of lethal prostate cancer that suppresses the androgen axis. Nat Med. 2018;24(12):1887–98. Epub 2018/11/28. doi: 10.1038/s41591-018-0241-1. PubMed PMID: 30478421; PMCID: PMC6614557.https://www.ncbi.nlm.nih.gov/pubmed/30478421

34. Aytes A, Mitrofanova A, Lefebvre C, Alvarez MJ, Castillo-Martin M, Zheng T, Eastham JA, Gopalan A, Pienta KJ, Shen MM, Califano A, Abate-Shen C. Cross-species regulatory network analysis identifies a synergistic interaction between FOXM1 and CENPF that drives prostate cancer malignancy. Cancer Cell. 2014;25(5):638–51. Epub 2014/05/16. doi: 10.1016/j.ccr.2014.03.017. PubMed PMID: 24823640; PMCID: PMC4051317.https://www.ncbi.nlm.nih.gov/pubmed/24823640

35. Vidal SJ, Rodriguez-Bravo V, Quinn SA, Rodriguez-Barrueco R, Lujambio A, Williams E, Sun X, de la Iglesia-Vicente J, Lee A, Readhead B, Chen X, Galsky M, Esteve B, Petrylak DP, Dudley JT, Rabadan R, Silva JM, Hoshida Y, Lowe SW, Cordon-Cardo C, Domingo-Domenech J. A targetable GATA2-IGF2 axis confers aggressiveness in lethal prostate cancer. Cancer Cell. 2015;27(2):223–39. Epub 2015/02/12. doi: 10.1016/j.ccell.2014.11.013. PubMed PMID: 25670080; PMCID: PMC4356948.https://www.ncbi.nlm.nih.gov/pubmed/25670080

36. Santasusagna S, Zhu S, Jawalagatti V, Carceles-Cordon M, Ertel A, Garcia-Longarte S, Song WM, Fujiwara N, Li P, Mendizabal I, Petrylak DP, Kelly WK, Reddy EP, Wang L, Schiewer MJ, Lujambio A, Karnes J, Knudsen KE, Cordon-Cardo C, Dong H, Huang H, Carracedo A, Hoshida Y, Rodriguez-Bravo V, Domingo-Domenech J. Master Transcription Factor Reprogramming Unleashes Selective Translation Promoting Castration Resistance and Immune Evasion in Lethal Prostate Cancer. Cancer Discov. 2023;13(12):2584–609. Epub 2023/09/07. doi: 10.1158/2159-8290.CD-23-0306. PubMed PMID: 37676710; PMCID: PMC10714140.https://www.ncbi.nlm.nih.gov/pubmed/37676710

37. Ichihara-Tanaka K, Kadomatsu K, Kishida S. Temporally and Spatially Regulated Expression of the Linker Histone H1fx During Mouse Development. J Histochem Cytochem. 2017;65(9):513–30. Epub 2017/08/03. doi: 10.1369/0022155417723914. PubMed PMID: 28766996; PMCID: PMC5582669.https://www.ncbi.nlm.nih.gov/pubmed/28766996

38. Zhou F, Liu Q, Zhang L, Zhu Q, Wang S, Zhu K, Deng R, Liu Y, Yuan G, Wang X, Zhou L. Selective inhibition of CBP/p300 HAT by A-485 results in suppression of lipogenesis and hepatic gluconeogenesis. Cell Death Dis. 2020;11(9):745. Epub 2020/09/13. doi: 10.1038/s41419-020-02960-6. PubMed PMID: 32917859; PMCID: PMC7486386.https://www.ncbi.nlm.nih.gov/pubmed/32917859

39. Martins VF, LaBarge SA, Stanley A, Svensson K, Hung CW, Keinan O, Ciaraldi TP, Banoian D, Park JE, Ha C, Hetrick B, Meyer GA, Philp A, David LL, Henry RR, Aslan JE, Saltiel AR, McCurdy CE, Schenk S. p300 or CBP is required for insulin-stimulated glucose uptake in skeletal muscle and adipocytes. JCI Insight. 2022;7(1). Epub 2021/11/24. doi: 10.1172/jci.insight.141344. PubMed PMID: 34813504; PMCID: PMC8765050.https://www.ncbi.nlm.nih.gov/pubmed/34813504

40. Cui A, Xue Y, Su W, Lin J, Liu Y, Cai G, Wan Q, Jiang Y, Ding D, Zheng Z, Wei S, Li W, Shen J, Wen J, Huang M, Zhao J, Zhang X, Zhao Y, Li H, Ying H, Zhang H, Bi Y, Chen Y, Xu A, Xu Y, Li Y. Glucose regulation of adipose tissue browning by CBP/p300- and HDAC3-mediated reversible acetylation of CREBZF. Proc Natl Acad Sci U S A. 2024;121(16):e2318935121. Epub 2024/04/08. doi: 10.1073/pnas.2318935121. PubMed PMID: 38588421; PMCID: PMC11032498.https://www.ncbi.nlm.nih.gov/pubmed/38588421

41. Wang S, Li T, Sheng C, Tan J, Yang Y, Ma X, Liu Y, Wei R, Zhou F, Zhou L, Wang X. CBP/p300 is critical for the expansion and maintenance of functional pancreatic alpha cell mass. Nat Commun. 2026;17(1). Epub 2026/04/12. doi: 10.1038/s41467-026-71499-5. PubMed PMID: 41965878; PMCID: PMC13247099.https://www.ncbi.nlm.nih.gov/pubmed/41965878

42. Beaubier N, Bontrager M, Huether R, Igartua C, Lau D, Tell R, Bobe AM, Bush S, Chang AL, Hoskinson DC, Khan AA, Kudalkar E, Leibowitz BD, Lozachmeur A, Michuda J, Parsons J, Perera JF, Salahudeen A, Shah KP, Taxter T, Zhu W, White KP. Integrated genomic profiling expands clinical options for patients with cancer. Nat Biotechnol. 2019;37(11):1351–60. Epub 20190930. doi: 10.1038/s41587-019-0259-z. PubMed PMID: 31570899.https://www.ncbi.nlm.nih.gov/pubmed/31570899

43. Langfelder P, Horvath S. WGCNA: an R package for weighted correlation network analysis. BMC Bioinformatics. 2008;9:559. Epub 2008/12/31. doi: 10.1186/1471-2105-9-559. PubMed PMID: 19114008; PMCID: PMC2631488.https://www.ncbi.nlm.nih.gov/pubmed/19114008

44. Ware KE, Somarelli JA, Schaeffer D, Li J, Zhang T, Park S, Patierno SR, Freedman J, Foo WC, Garcia-Blanco MA, Armstrong AJ. Snail promotes resistance to enzalutamide through regulation of androgen receptor activity in prostate cancer. Oncotarget. 2016;7(31):50507–21. Epub 2016/07/14. doi: 10.18632/oncotarget.10476. PubMed PMID: 27409172; PMCID: PMC5226599.https://www.ncbi.nlm.nih.gov/pubmed/27409172

