## Supplemental Figure 1 for "Integrated real-world and translational studies nominate CREB binding protein/EP300 inhibition to overcome androgen receptor pathway modulator resistance in men with prostate cancer"

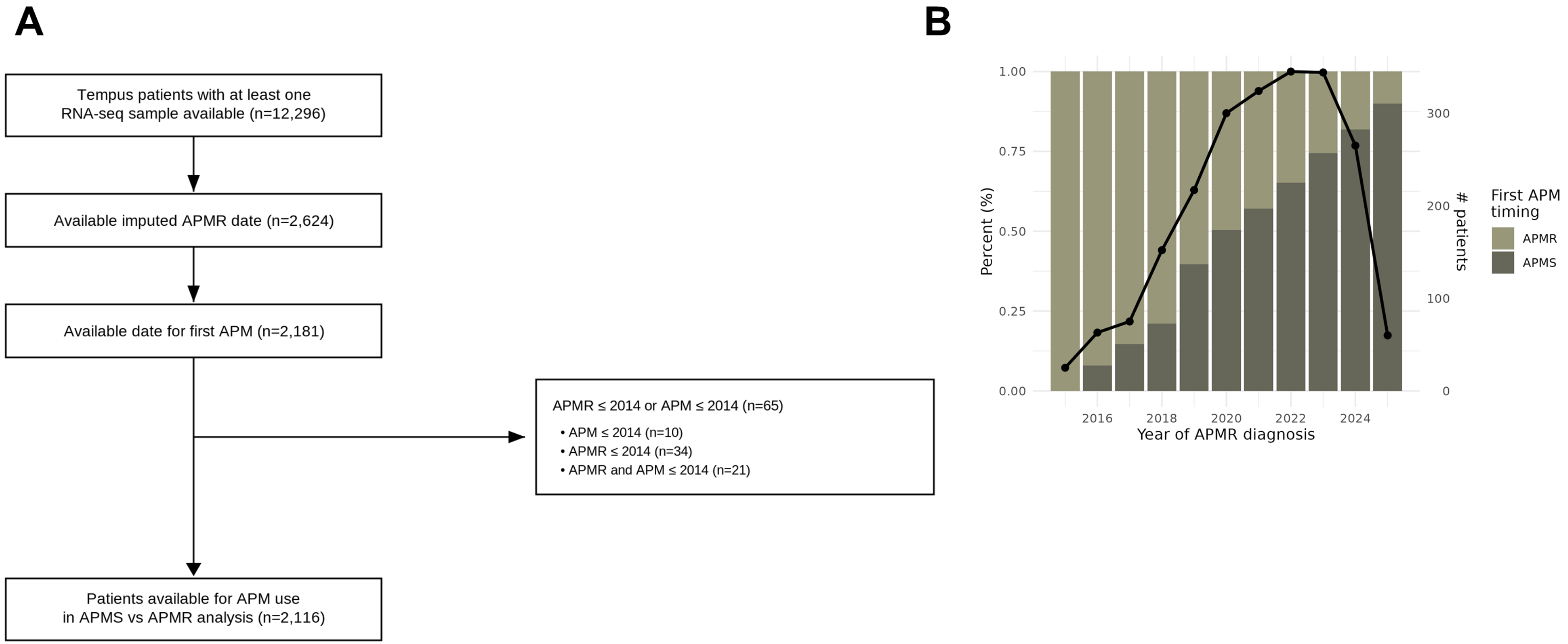
A
Tempus patients with at least one RNA-seq sample available (n=12,296)
Available imputed APMR date (n=2,624)
Available date for first APM (n=2,181)
APMR ≤ 2014 or APM ≤ 2014 (n=65)
• APM ≤ 2014 (n=10)
• APMR ≤ 2014 (n=34)
• APMR and APM ≤ 2014 (n=21)
Patients available for APM use in APMS vs APMR analysis (n=2,116)
B
Percent (%)
1.00
0.75
0.50
0.25
0.00
### patients
300
200
100
0
2016
2018
2020
2022
2024
Year of APMR diagnosis
First APM timing
APMR
APMS
