## Supplemental Figure 2 for "Integrated real-world and translational studies nominate CREB binding protein/EP300 inhibition to overcome androgen receptor pathway modulator resistance in men with prostate cancer"

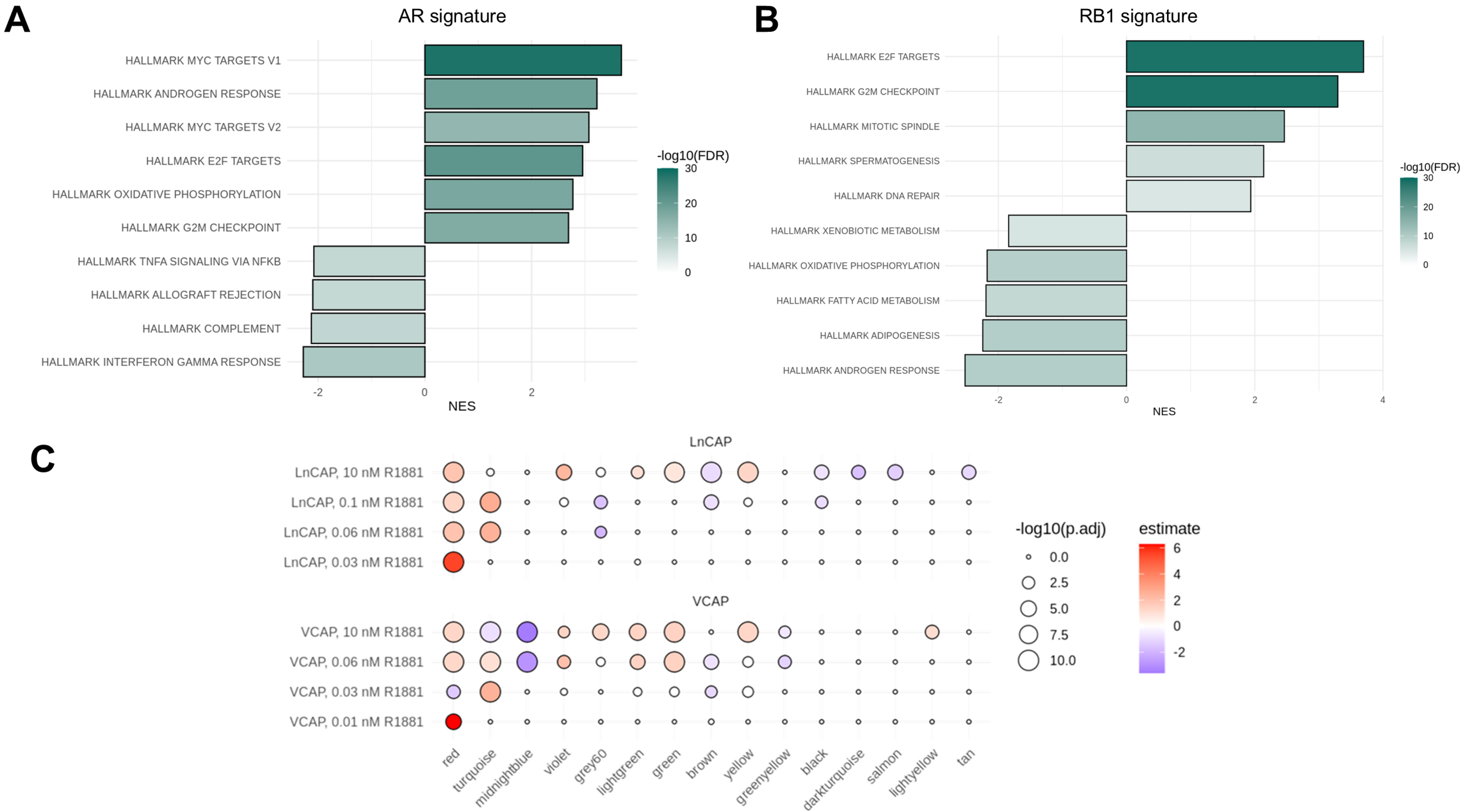
A
AR signature
HALLMARK MYC TARGETS V1
HALLMARK ANDROGEN RESPONSE
HALLMARK MYC TARGETS V2
HALLMARK E2F TARGETS
HALLMARK OXIDATIVE PHOSPHORYLATION
HALLMARK G2M CHECKPOINT
HALLMARK TNFA SIGNALING VIA NFKB
HALLMARK ALLOGRAFT REJECTION
HALLMARK COMPLEMENT
HALLMARK INTERFERON GAMMA RESPONSE
-2
0
2
NES
-log10(FDR)
30
20
10
0
B
RB1 signature
HALLMARK E2F TARGETS
HALLMARK G2M CHECKPOINT
HALLMARK MITOTIC SPINDLE
HALLMARK SPERMATOGENESIS
HALLMARK DNA REPAIR
HALLMARK XENOBIOTIC METABOLISM
HALLMARK OXIDATIVE PHOSPHORYLATION
HALLMARK FATTY ACID METABOLISM
HALLMARK ADIPOGENESIS
HALLMARK ANDROGEN RESPONSE
-2
0
2
4
NES
-log10(FDR)
30
20
10
0
C
LnCAP
LnCAP, 10 nM R1881
LnCAP, 0.1 nM R1881
LnCAP, 0.06 nM R1881
LnCAP, 0.03 nM R1881
VCAP
VCAP, 10 nM R1881
VCAP, 0.06 nM R1881
VCAP, 0.03 nM R1881
VCAP, 0.01 nM R1881
red
turquoise
midnightblue
violet
grey60
lightgreen
green
brown
yellow
greenyellow
black
darkturquoise
salmon
lightyellow
tan
-log10(p.adj)
0.0
2.5
5.0
7.5
10.0
estimate
6
4
2
0
-2
