## Supplemental Figure 3 for "Integrated real-world and translational studies nominate CREB binding protein/EP300 inhibition to overcome androgen receptor pathway modulator resistance in men with prostate cancer"

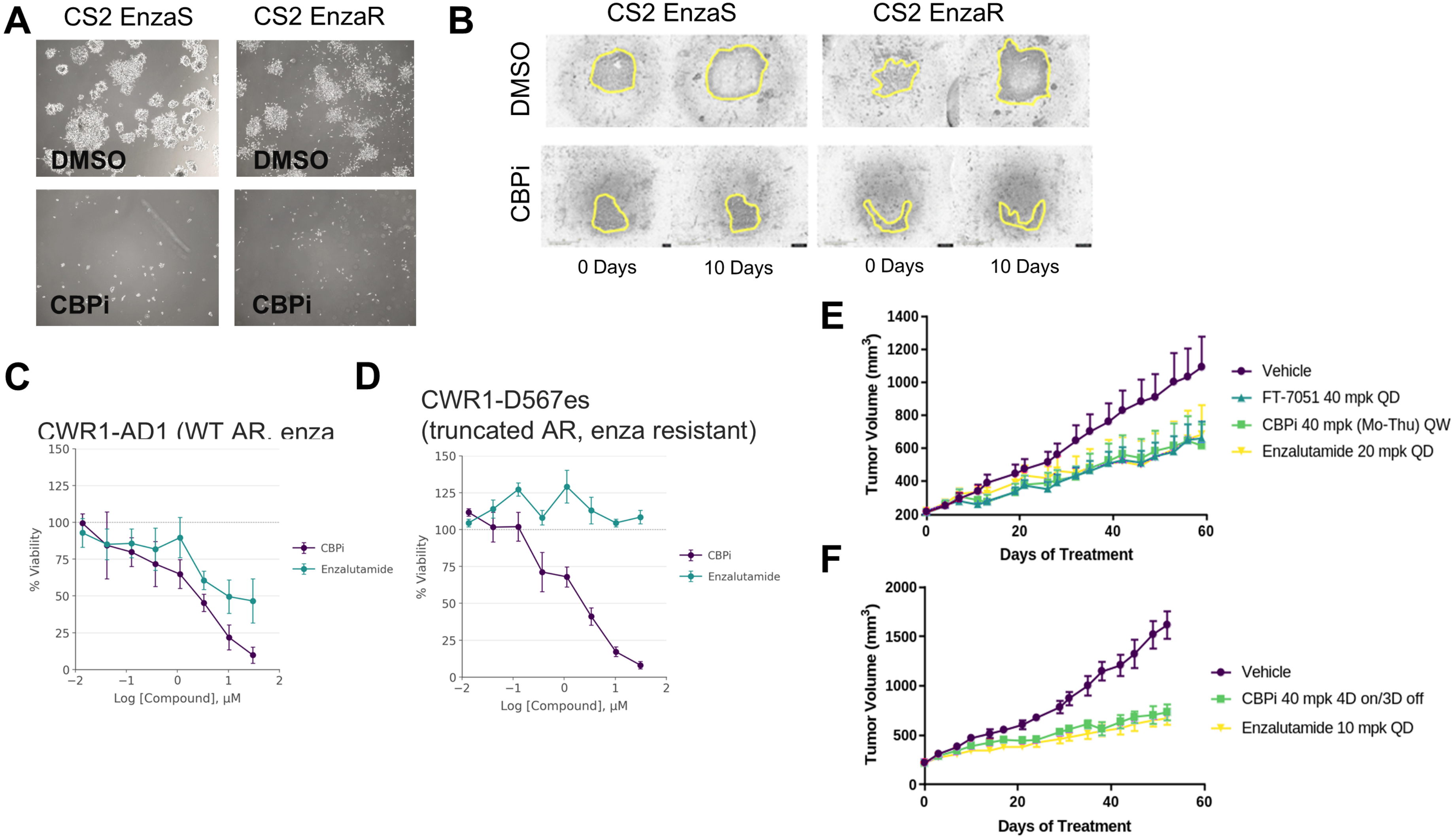
A
CS2 EnzaS
CS2 EnzaR
DMSO
DMSO
CBPi
CBPi
B
CS2 EnzaS
CS2 EnzaR
DMSO
CBPi
0 Days
10 Days
0 Days
10 Days
C
CWR1-AD1 (WT AR, enza
% Viability
Log [Compound], μM
CBPi
Enzalutamide
D
CWR1-D567es
(truncated AR, enza resistant)
% Viability
Log [Compound], μM
CBPi
Enzalutamide
E
Tumor Volume (mm$^3$)
Days of Treatment
Vehicle
FT-7051 40 mpk QD
CBPi 40 mpk (Mo-Thu) QW
Enzalutamide 20 mpk QD
F
Tumor Volume (mm$^3$)
Days of Treatment
Vehicle
CBPi 40 mpk 4D on/3D off
Enzalutamide 10 mpk QD
